# Targeted metagenomic sequencing for detection of vertebrate viruses in wastewater for public health surveillance

**DOI:** 10.1101/2023.03.14.23287251

**Authors:** Camille McCall, Ryan A. Leo Elworth, Kristine M. Wylie, Todd N. Wylie, Katherine Dyson, Ryan Doughty, Todd J. Treangen, Loren Hopkins, Katherine Ensor, Lauren B. Stadler

**Affiliations:** Department of Civil and Environmental Engineering, Rice University, Houston, TX, 77005; Department of Computer Science, Rice University, Houston, TX, 77005; Department of Pediatrics, Washington University School of Medicine, St. Louis, Missouri 63110; Houston Health Department, 8000 N. Stadium Dr., Houston, TX, 77054; Department of Statistics, Rice University, Houston, TX, 77005

**Keywords:** Wastewater-based epidemiology, metagenomic shotgun sequencing, viruses, capture sequencing, astroviruses

## Abstract

Viruses of concern for quantitative wastewater monitoring are usually selected as a result of an outbreak and subsequent detection in wastewater. However, targeted metagenomics could proactively identify viruses of concern when used as an initial screening tool. To evaluate the utility of targeted metagenomics for wastewater screening, we used ViroCap, a panel of probes designed to target all known vertebrate viruses. Untreated wastewater was collected from wastewater treatment plants (WWTPs) and building-level manholes associated with vulnerable populations in Houston, TX. We evaluated differences in vertebrate virus detection between WWTP and building-level samples, classified human viruses in wastewater, and performed phylogenetic analysis on astrovirus sequencing reads to evaluate targeted metagenomics for subspecies level classification. Vertebrate viruses varied widely across building-level samples. Rarely detected and abundant viruses were identified in WWTP and building-level samples, including enteric, respiratory, and bloodborne viruses. Furthermore, full length genomes were assembled from astrovirus reads and two human astrovirus serotypes were classified in wastewater samples. This study demonstrates the utility of targeted metagenomics as an initial screening step for public health surveillance.

**Synopsis:** This work demonstrates the utility of targeted metagenomic shotgun sequencing to screen for human viruses in centralized and building-level wastewater samples.

**For Table of Contents Only:** 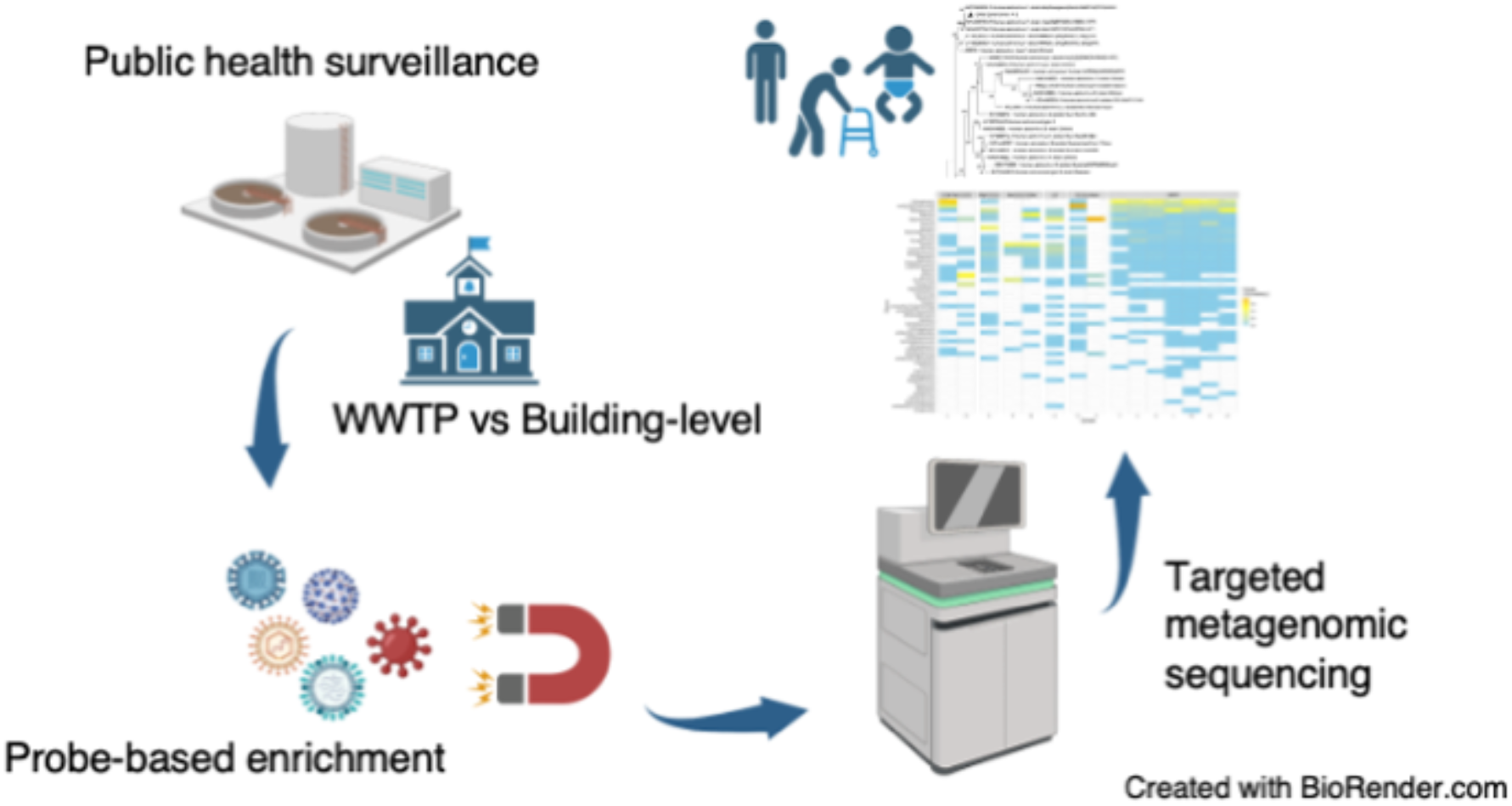

## 1. Introduction

Approximately 75% of the US population is connected to a municipal wastewater treatment facility (Westerhoff et al., 2015), which collects sewage containing a diverse range of commensal and pathogenic organisms. Human vertebrate viruses excreted through feces, urine, skin, saliva, and blood can be detected in wastewater to gather insights into viral diseases circulating within a population. Previous studies have associated viral presence in wastewater with outbreaks in the corresponding community (Bisseux et al., 2018; Farkas et al., 2018; Fernandez-Cassi et al., 2021; Galani et al., 2022; Hellmér et al., 2014; McCall et al., 2021; Peccia et al., 2020; Prevost et al., 2015; Zhao et al., 2022). More recently, the CDC and public health departments have deemed wastewater monitoring as an integral public health surveillance tool for early detection and mitigation of COVID-19 (Kirby et al., 2021). Viruses of concern for long-term quantitative wastewater monitoring are usually selected as a result of clinical presence and subsequent detection in municipal wastewater. Consequently, viruses that have significant incubation periods, subclinical presentations, or are highly infectious may reach thousands of people before healthcare facilities are notified and the virus is incorporated into the wastewater monitoring framework. The integration of sensitive, unbiased methods into the existing framework is needed to proactively identify and select viruses of concern for public health surveillance.

Untargeted metagenomic analysis provides a relatively unbiased approach to detecting viruses in wastewater. Several studies have explored the presence of viral pathogens in wastewater using metagenomics (Aw et al., 2014; Bibby and Peccia, 2013; Cantalupo et al., 2011; Fernandez-Cassi et al., 2020; Martínez-Puchol et al., 2020; McCall et al., 2020; O’Brien et al., 2017). Metagenomics has identified novel viruses, causative agents to disease outbreaks, and differences in viral diversity between geographical regions (Matus et al., 2019; Newton et al., 2015; Ng et al., 2012; Xu et al., 2011). Most wastewater viral metagenomic studies incorporate a sample purification step, particularly, a concentration step, prior to processing samples for sequencing. Concentration is important because human viruses are present at a low abundance in wastewater and wastewater contains inhibitors that can interfere with the detection of such viruses. Membrane filtration is a common concentration method used, either as a stand-alone method or in conjunction with other methods, in wastewater metagenomic studies (McCall et al. 2019). Although concentration can improve virus detection, it can impose bias during sample processing depending on several factors such as the method used and virus morphology (Hjelmsø et al., 2017, Girón-Guzmán et al., 2023). More studies are needed to explore the impact of concentration biases on virus detection in metagenomic datasets.

More recently, targeted metagenomics has been introduced to increase the sensitivity of human virus detection in wastewater. One such example of targeted metagenomic is in-solution hybridization capture sequencing, which uses probes and magnetic bead technology to enrich for a comprehensive set of vertebrate virus genomes (Wylie et al., 2015). This is especially important for low abundance viruses such as human viruses that may go undetected in wastewater metagenomes, which harbor a host of larger, more dominant viruses such as plant viruses and bacteriophages. Furthermore, a previous study that compared targeted metagenomic sequencing and amplicon deep sequencing was able to identify species-level information of several different viruses, including enteroviruses, using both methods (Martínez-Puchol et al., 2020). This highlights the potential for targeted metagenomic sequencing to identify species and subspecies of important viral pathogens.

Wastewater sampling for metagenomic analysis is commonly carried out at a wastewater treatment plant (WWTP). Samples collected from WWTPs can provide a snapshot of community health across large populations, identify prevalent viruses circulating within a city or county, and provide a means to accessible and convenient sampling locations. Although sampling at a WWTP is useful for understanding population-wide disease burdens, viral disease burdens often vary between different localized populations and demographic groups. A recent study found neighborhood-level wastewater to contain biomarkers that are correlated with certain socio-economic and sociodemographic factors (Choi et al., 2019). The pooling of wastewater in sewer networks can create a homogenized view of viral presence, highlighting only viruses that are most abundant across the population as a whole. Therefore, local outbreaks (e.g., neighborhood, building-level) may go undetected in WWTP samples. Building-level sampling provides more geographically-resolved data, higher specificity, and identification of viruses that are prevalent among high-risk groups — specifically, children, elderly, incarcerated individuals, and the homeless. This information can be used to select viruses for long-term quantitative wastewater monitoring according to a community’s need and tailor intervention strategies to specific populations. Few studies have implemented metagenomic sequencing for virus detection at the building-level (Spurbeck et al., 2021) and to our knowledge no studies have implemented viral metagenomics on the wastewater of vulnerable populations such as those inhabiting jails or homeless shelters.

This study evaluates targeted metagenomics via probe-based capture enrichment prior to sequencing to facilitate the detection of vertebrate viruses for public health surveillance. Untreated wastewater samples were collected from 7 WWTPs and 8 building-level manholes in Houston, TX associated with high-risk populations, including a jail, homeless shelters, child care centers, a high school, and nursing homes. Targeted and untargeted metagenomic datasets were compared to quantify differences in virus detection between sequencing methods. Targeted and untargeted samples underwent the same sample processing methods, specifically electronegative membrane filtration (MF), prior to probe-based capture enrichment (i.e., targeted/MF and untargeted/MF).

Additionally, we compared vertebrate virus detection using targeted metagenomic shotgun sequencing in samples concentrated using electronegative membrane filtration (targeted/MF) and samples that underwent direct extraction (DE) (targeted/DE) to evaluate the impact of concentration on virus detection. Of note, because this study focuses on the impact of enrichment methods on virus detection in metagenomic data, untargeted/DE versus untargeted/MF and targeted/DE versus untargeted/DE were not considered. Vertebrate virus detection was then compared between WWTP and building-level wastewater samples for metagenomic datasets that were concentrated and subsequently targeted using capture sequencing. A discussion of the human viruses that were classified in all concentrated/targeted wastewater samples is also provided. Species-level sequencing reads were further processed to assess the potential for targeted metagenomics to identify viruses at the subspecies level in wastewater samples. This study focused on astroviruses since they are abundant in wastewater and an important pathogen among children and elderly.

## 2. Methods

### 2.1. Study areas and wastewater sample collection

Centralized wastewater samples were collected from the influent of 7 WWTPs (I-O) within Houston, Texas covering 7 different sewersheds. In total, the WWTPs collect wastewater from approximately 86% of the city’s population. Building-level samples were collected from 8 manhole locations (A-H) with each one receiving untreated wastewater directly from their respective facilities (child care center (A, B), high school (C), homeless shelter (F, G), jail (H), nursing home (D, E)). All samples were collected from 24-hour composite samplers. Wastewater samples were aliquoted into 250- or 500-mL sterile Nalgene bottles, and transported on ice to the Rice University NEST lab for further processing.

### 2.2. Sample concentration and extraction

Samples undergoing filtration were concentrated within 24 hours of arrival. Concentration was performed in duplicate for each wastewater sample by first aliquoting roughly 50 mL of sample into 50mL conical tubes and centrifuging for 10 minutes at 4,100 g and 4°C to remove solids. Samples were then filtered using electronegative membrane HA filters (HAWG047S6, Millipore Sigma). Samples filters were transferred to 2 mL bead mill tubes followed by bead beating as previously described (LaTurner et al., 2021) and briefly explained below. To evaluate the difference between targeted and untargeted sequencing, and membrane filtration and direct extraction on vertebrate virus detection in metagenomic datasets, WWTP samples M and O were divided into two additional replicates each. One out of two of the additional replicates for each WWTP sample were filtered as described above, and the remaining replicate was processed using direct extraction. For direction extraction, each 50 mL replicate was well mixed and 300 uL of sample was transferred to a bead tube. For both filtered and unfiltered samples, bead beating tubes contained 0.1 mm glass bead and 1 mL of lysis buffer. Bead beating was carried out with XX instrument and YY conditions, and total nucleic acid was extracted on a chemagic 360 automated platform using the Viral DNA/RNA 300 Kit H96 (CMG-1433, PerkinElmer) following the manufacturer’s protocol.

### 2.3. Two-strand cDNA synthesis and probe-based capture sequencing

To capture both DNA and RNA viruses, total nucleic acid from each sample was subject to double stranded cDNA synthesis according to (Wang et al., 2003) with some modifications. Specifically, the random primer was modified to contain only the nanomer region of the sequence to eliminate the presence of Primer B nucleotides in sequencing reads. Two-strand cDNA synthesis for each sample was carried out in a 30 uL reaction mixture containing 40 uM of random primer and 5 uL of nucleic acid extract. The complete protocol, including reaction mixture components and thermocycling conditions, is detailed in Appendix 1 of the supplementary information. Following cDNA synthesis, samples were shipped on dry ice to the Genome Technology Access Center at McDonnell Genome Institute at Washington University in Saint Louis, MO for indexed sequencing library construction with the Kapa HyperPrep kit (Roche). For libraries that were enriched with ViroCap, sequencing libraries were pooled and combined with the ViroCap panel, which consists of probes designed to target all known DNA and RNA vertebrate viruses (Wylie et al., 2015). Enrichment was carried out according to the manufacturer’s instructions (Roche). The two additional replicates that were filtered for WWTPs M and O were sequencing directly without ViroCap enrichment. Illumina sequencing for targeted and untargeted samples was performed on a NovaSeq platform generating 150 bp paired-end reads. Sequences were demultiplexed using the index sequences from each sample’s sequencing library. The raw sequence reads used in this study can be found in the Sequence Read Archive (SRA) under BioProject ID: PRJNA943189.

### 2.4. Metagenomic and phylogenetic analysis

Sequencing reads were analyzed using the ViroMatch pipeline (Wylie and Wylie, 2021). In summary, reads were quality filtered, host screened, and aligned to virus-specific reference genomes obtained from NCBI GenBank. Multistep alignment was performed using BWA-MEM for nucleotide mapping and DIAMOND for translated nucleotide mapping. If a virus obtained assigned reads for both alignment strategies, the assigned reads were combined to get the total number of assigned reads for that virus. A threshold of 0.1% of the total assigned reads across all samples for each virus detected was subtracted as background to account for sequencing crosstalk observed in pooled sequencing libraries (Wylie et al., 2018).

Reads aligning to mamastrovirus (MAstV) genus were collected and further processed to assess the genetic diversity of human astroviruses in wastewater samples. MAstV reads per sample were combined into individual R1 and R2 FASTQ files based on their species-level classification and assembled into contigs using IDBA-UD. IDBA-UD is useful for strain-level analysis due to its high accuracy and genome recovery (Sutton et al., 2019). Contigs with sequencing lengths comparable to the complete genome of the appropriate MAstV were considered. Putative whole genome contigs were aligned against their respective sequencing reads to determine the percent coverage and average read depth per contig using BWA-MEM under default conditions. Contig alignment with BWA-MEM was visually inspected using the Integrative Genomics Viewer (IGV) to evaluate the quality of each contig. Potential misassembled contigs were amended, if feasible, before inclusion in multiple sequence alignment (MSA).

In our samples, MAstV-1 was the only astrovirus species for which we obtained contigs comparable to the length of its whole genome (∼6.7 kbp). Sequence assembly generated a total of 3 MAstV-1 contigs from child care center A (n=2) and WWTP M (n=1). Child care center A-1 contig and the WWTP M contig were divided at 5.8 kbp and 4.4 kbp, respectively, to remove gaps in read coverage. Therefore, child care center A-1 and WWTP M contigs were considered near complete genome contigs. Contig coverage results and lengths are provided in Table S1 and Figures S1-S3.

Homology and phylogenetic analyses were carried out with 38 complete MAstV reference genomes obtained from NCBI GenBank, representing each human MAstV-1 serotype, and used as references for MSA and phylogenetic analysis. MSA was performed using MUSCLE. All alignment files were forwarded to Fasttree 2.1.11 to create maximum likelihood trees using the generalized time-reversible model and a bootstrap test with 1000 replicates. The remainder of the parameters were run under default conditions. Tree files were generated in Newick format and visualized using MEGA 11.0.

### 2.5. Statistical Analysis

All statistical analyses were performed in R (R Core Team, 2022). Prcomp was used for the PCoA. Welch’s student t-test was used to evaluate the difference between samples after verifying the normality of the datasets using the Shapiro-Wilk test and qq plots. A p-value < 0.05 was considered statistically significant.

## 3. Results and Discussion

Metagenomic shotgun sequencing enriched by ViroCap probe-based hybridization capture was used to screen WWTP and building-level wastewater for vertebrate viruses including human pathogens. Targeted sequencing was performed on 8 building-level samples (A-H) and 7 WWTP samples (I-O). Illumina sequencing resulted in a total of 171 Gb (gigabases) or approximately 1.2 billion reads. A total of 1.1 million (0.1%) reads were assigned to viral taxonomic groups via the ViroMatch pipeline. After applying the contamination threshold, the number of viral affiliated reads reduced by 0.09%.

### 3.1. Targeted metagenomics with electronegative membrane filtration increased vertebrate virus detection in wastewater samples when compared to untargeted and direction extraction methods

Wastewater samples M and O were divided into two additional replicates each to evaluate the impact of electronegative membrane filtration and targeted sequencing on vertebrate virus detection in sequencing reads. Targeted metagenomic sequencing coupled with membrane filtration (targeted/MF) resulted in a 35% (n=20) and 67% (n=18) increase in the number of unique vertebrate viral families detected in the M and O WWTPs, respectively, as compared to the untargeted/MF samples for WWTPs M (n=13) and O (n=6) (Figure S4). All viral families that were detected in the untargeted/MF sample were also detected in the targeted/MF sample for both WWTPs. The number of reads per vertebrate viral family generally increased in targeted samples with the exception of Poxviridae, which decreased in both wastewater samples (Table S2). Poxviridae family contains viruses that infect vertebrates as well as viruses that infect Arthropods, invertebrate animals. Hence, the decrease in Poxviridae reads was due to a decrease in viruses with invertebrate hosts (data not shown), which are not included in the ViroCap panel to date. Additional viral targets can be added to the panel if invertebrate viruses are of interest. Findings here suggest that targeted metagenomic sequencing with ViroCap increases detection and read counts of targeted viruses. This was consistent with other studies that used targeted metagenomics for virus detection (Martínez-Puchol et al., 2020; Paskey et al., 2019; Wylie et al., 2015).

Similarly, targeted/MF increased the number of vertebrate viral families detected by more than half in both M and O WWTP samples as compared to samples that were targeted and directly extracted (targeted/DE), 80% (n= 4) and 67% (n=6), respectively (Figure S4). Similarly, Segelhurst et al., 2022 used a tangential-flow filtration method to recover 53% more complete or near complete SARS-CoV-2 genomes from wastewater samples as compared to concentration using precipitation. Still, a previous study by Hjelmsø et al., 2017 evaluated the impact of polyethylene glycol precipitation (PEG), skimmed milk flocculation, glass wool filtration, and monolithic adsorption filtration on the sequencing of viruses in wastewater. They demonstrated that concentration methods significantly impacted the detection of viruses in metagenomic sequencing reads with PEG concentrated samples obtaining more viral-affiliated reads compared to the other methods. Although the application of the two above mentioned studies is different, it is clear that more work is needed to understand the impact of concentration methods on the presence of viruses in sequencing data. Concentration methods can also impose bias on viral recovery considering the multiple forms in which viruses can exist in wastewater (i.e., intact, non-intact) and differences in virus morphology (i.e., enveloped, nonenveloped) (Mondal et al., 2021). In fact, Giron-Guzman et al. 2023 found that the Enviro Wastewater TNA concentration kit was more effective at recovering non-enveloped viruses from wastewater as compared to an aluminum-based adsorption-precipitation method. The opposite was true for enveloped viruses (Girón-Guzmán et al., 2023). This makes direct extraction a plausible approach to circumvent such challenges. However, raw wastewater contains high concentrations of organic matter and other inhibitors that can interfere with the detection and sensitivity of later processes. This necessitates the addition of one or multiple purification steps to increase the chances of virus detection in these complex sample matrices. Additionally, since viruses are present at dilute concentrations in wastewater, direct extraction can result in a lower equivalent volume of viruses in the final nucleic acid extract. A previous study found the use of centrifugation, filtration and nuclease-treatment to yield the highest proportion of viral affiliated sequences as compared to using only one or two of the aforementioned methods (Hall et al., 2014). Further, LaTurner et al., 2021 demonstrated that HA filtration with bead beating yielded the highest concentrations of SARS-CoV-2 as compared to other concentration methods tested including direct extraction. In this study, the highest viral yield was obtained when pairing membrane filtration with targeted metagenomic sequencing using ViroCap. This combination of methods was therefore used for the subsequent analysis below.

### 3.2. Vertebrate virus profiles were more homogenous across WWTP as compared to building-level samples

Viruses associated with vertebrates and bacteria constituted the greatest proportion of reads across all base samples (Figure 1). The total number of viral affiliated reads per sample is listed in Table S4. The proportion of vertebrate viruses across wastewater samples range between 0.06–0.95 with an average of 0.49. The average was approximately 26–43% higher than vertebrate virus detection in untargeted metagenomic studies (Aw et al., 2014; Ng et al., 2012; O’Brien et al., 2017). The proportion of vertebrate viruses was relatively consistent across WWTPs as compared to building-level samples where host range noticeably varied. This fluctuation in virus hosts could be due to differences in wastewater composition, diurnal cycles, and/or human input. Building-level samples can be more sensitive to minor changes in community activity as compared to WWTP samples.

**Figure 1.**
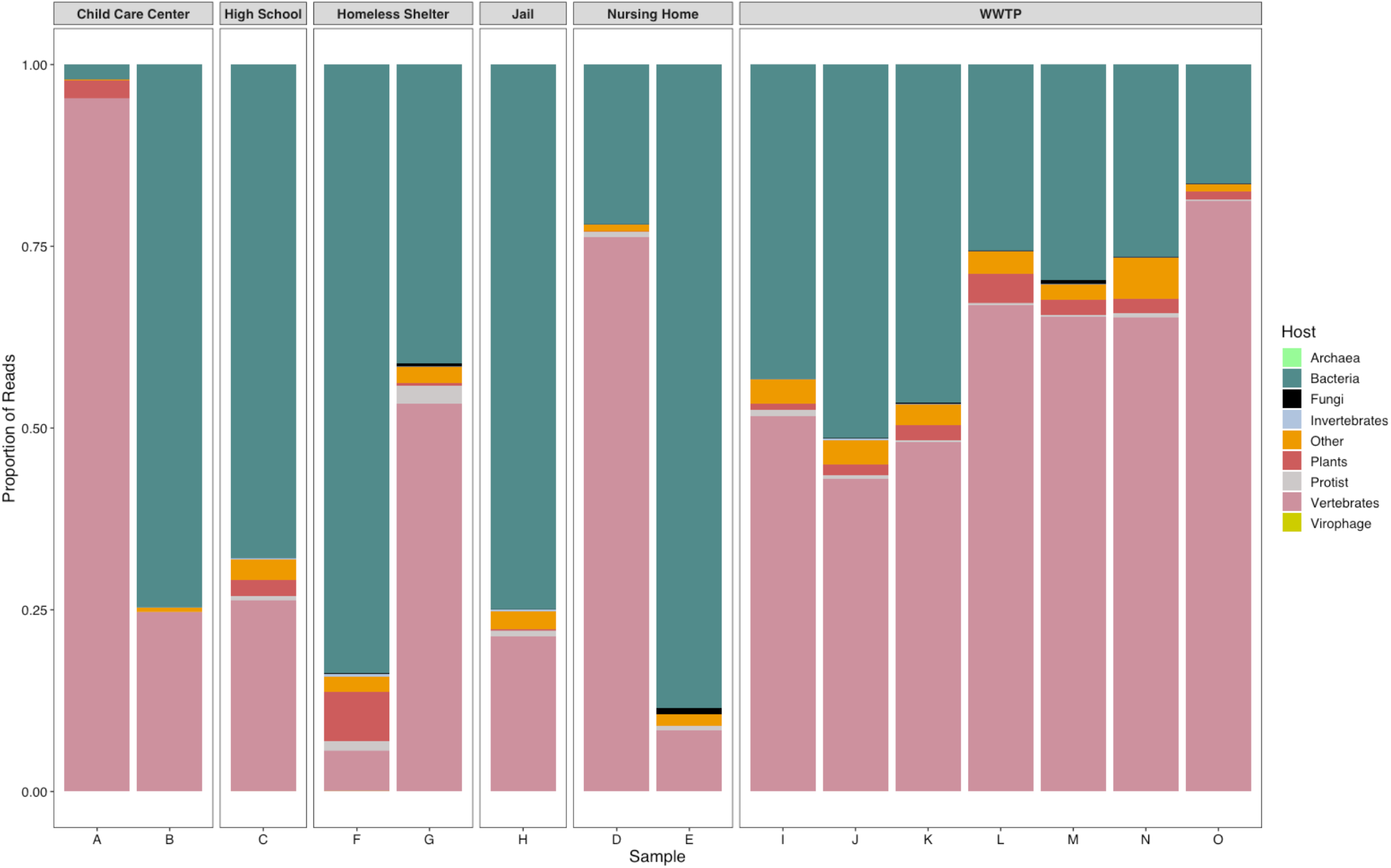
Proportion of hosts detected in WWTP (I-O) and building-level samples (A-H).

Twenty-four different vertebrate viral families were detected in wastewater samples with *Astroviridae* followed by *Adenoviridae*, and *Parvoviridae* obtaining the greatest number of hits across WWTPs. In total, *Astroviridae, Caliciviridae*, and *Picornaviridae* obtained the greatest number of hits in building-level samples (Figure 2). A principal component analysis was performed to determine the similarity between samples at the family taxonomic level (Figure 3). *Astroviridae, Coronaviridae*, and *Anelloviridae* viral families had the most influence when differentiating between samples. As expected, there were more similarities between WWTPs than building-level samples. The two nursing home samples were more similar to each other as compared to the pair of child care center and homeless shelter samples. More data is needed to sufficiently understand the differences in vertebrate virus detection between various communities.

**Figure 2.**
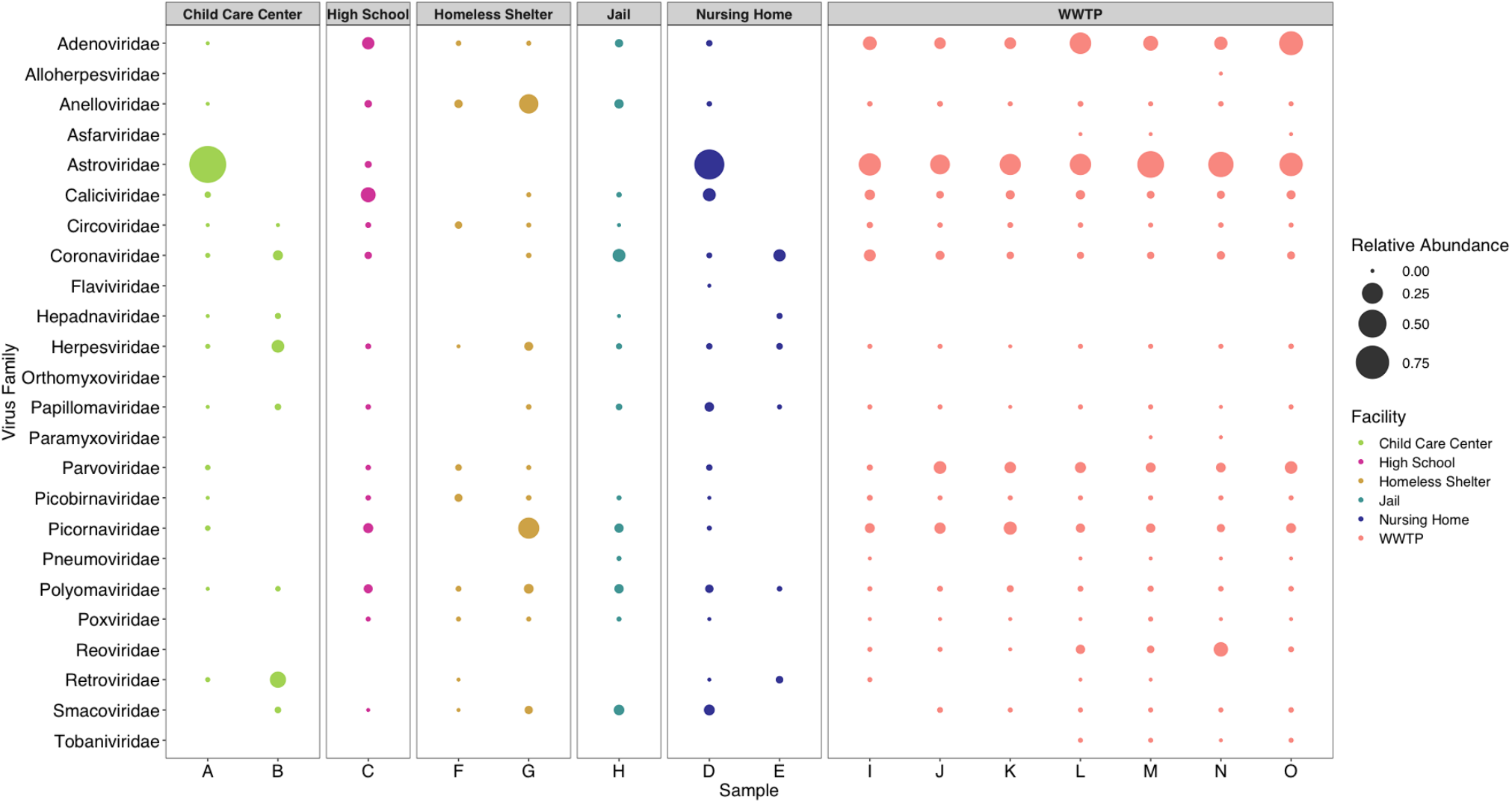
Classification of vertebrate virus families detected in WWTP and building-level samples.

**Figure 3.**
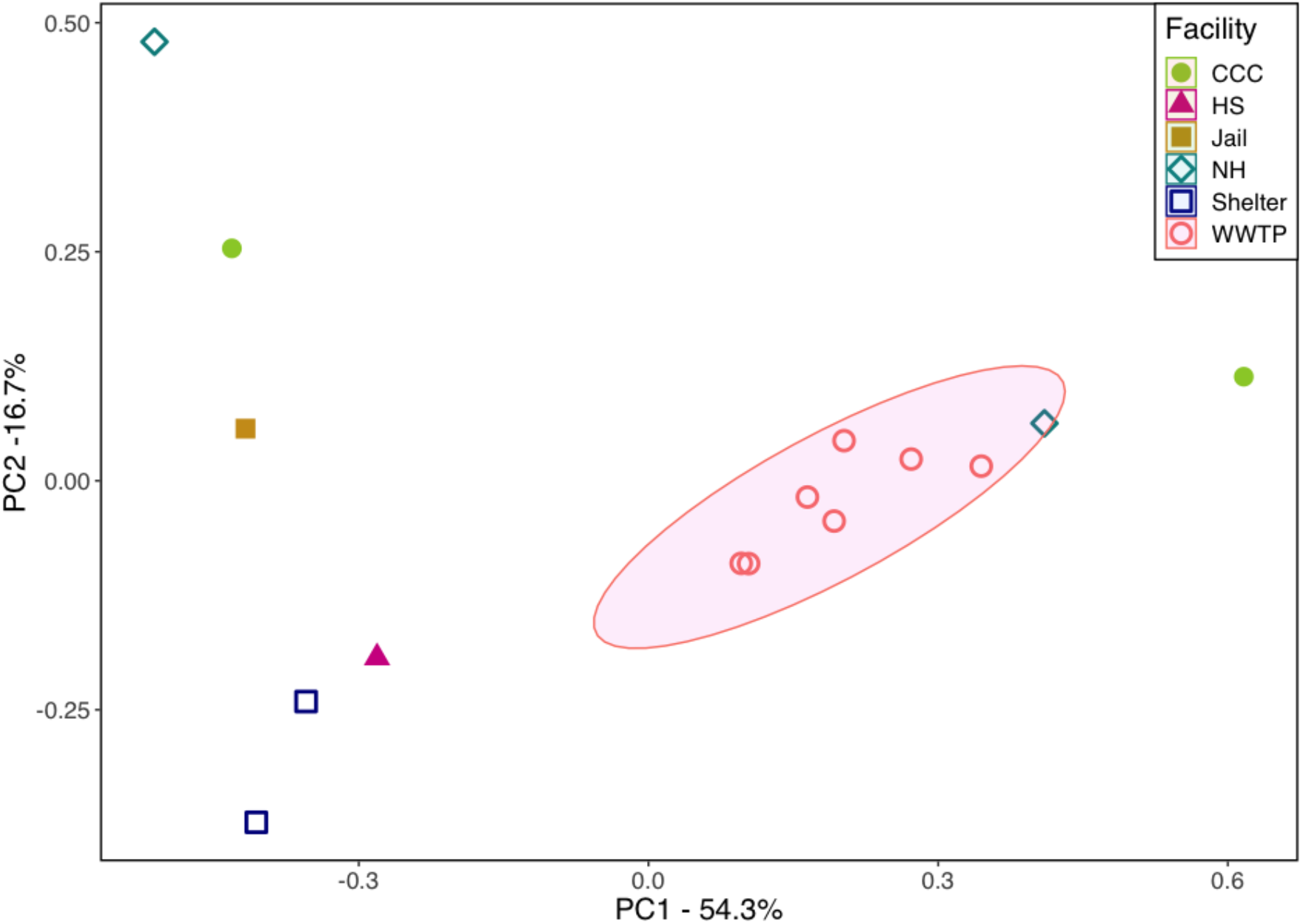
Principal component analysis (PCoA) of vertebrate viruses between sample types. Plot is generated at the family level. The wastewater samples are abbreviated as follows: child care center (CCC), high school (HS), jail (Jail), nursing home (NH), homeless shelter (Shelter), and wastewater treatment plant (WWTP). The polygon represents the 95% confidence level for a multivariate t-distribution.

### 3.3. Classification of human vertebrate viruses in Houston wastewater

Of the 24 vertebrate families detected, 20 of them were identified as human viral families containing 43 genera and 6 unclassified viral categories. Targeted metagenomic analysis identified respiratory, enteric, and other viral pathogens associated with blood, skin, and mucosal transmissions (Figure 4, Table S5). There were important distinctions in virus communities between WWTP and building-level samples. Generally, human viruses detected in building-level samples were less homogenous than WWTPs samples. Only 19–30% of viruses detected in the child care center, homeless shelter, or nursing home samples were also detected in the corresponding sample belonging to the same building type. Of the 49 viral groups identified, 4 were detected in building-level samples that went undetected in WWTP samples. Interestingly, the 4 viral groups that were not detected in the WWTP samples are non-enteric viruses, with orthohepadnavirus being rarely detected in wastewater samples (Table S5). Factors influencing virus detection in centralized WWTPs include their shedding patterns in human excrements, persistence in wastewater, dilution, and city-wide prevalence, as compared to localized occurrences. Since enteric viruses tend to dominate wastewater samples, non-enteric viruses can be challenging to detect, especially in large-scale WWTP samples. Findings here suggest that sampling at the building-level can increase the potential to identify viruses less commonly found in wastewater. Conversely, there were 12 viral groups detected in the WWTP samples that were not present in building-level samples. These viruses include both non-enteric and enteric viruses (Table S5). Since the WWTP samples contain wastewater collected from approximately 86% of the city’s population, it is likely that other populations within the sewershed contributed to the signal observed.

**Figure 4.**
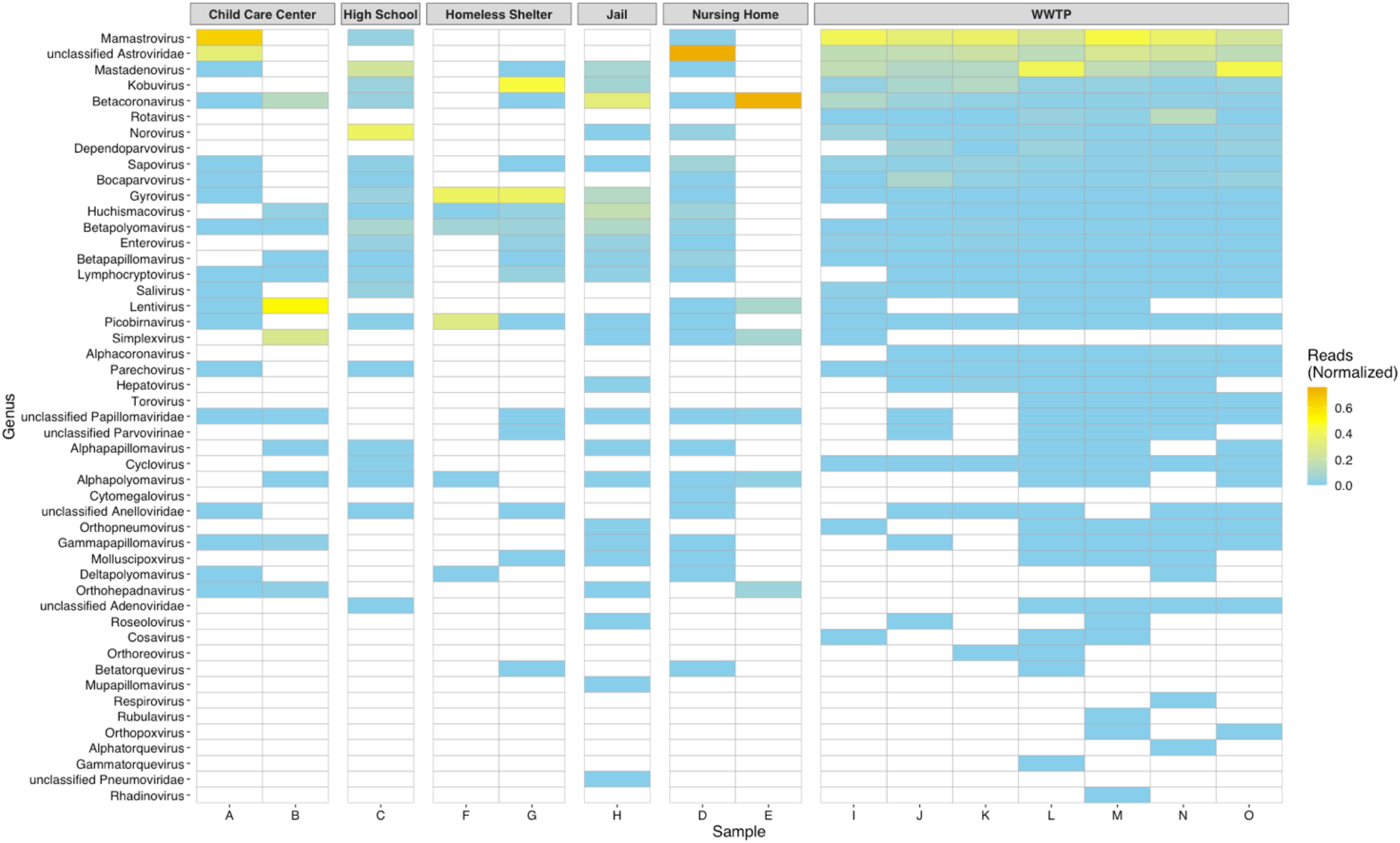
Heatmap of human genus virus diversity and normalized abundance in each sample. The normalized abundance was calculated by dividing the number of reads assigned to each genus by the total number of human viral-assigned reads in the corresponding samples. White cells indicate absence of associated virus in the sample. Genera are in descending order according to abundance.

Enteric followed by respiratory viruses collectively constituted the greatest proportion of reads in wastewater samples. Enteric pathogens generally cause gastrointestinal illnesses and are shed in significant concentrations in stool making them abundant in wastewater (Haramoto et al., 2018). Mamastrovirus (MAstV) and unclassified *Astroviridae* obtained the greatest number of hits among human viral reads. MAstV is a genus in the *Astroviridae* family infecting mammals. There are currently 19 species in the MAstV genus (MAstV-1-19) that are recognized by the International Committee on Taxonomy of Viruses (ICTV). Human viruses are classified within the following species: MAstV-1 (HAstV-1-8), MAstV-6 (MLB1-3), MAstV-8 (VA2/HMO-A, VA4), and MAstV-9 (VA1/HMO-C, VA3/HMO-B). HAstV-1-8 are considered classic HAstVs while the remaining serotypes are considered novel HAstVs (Donato and Vijaykrishna, 2017). In this study, MAstV-1, -6, and unclassified HAstVs obtained the majority of HAstV reads. Astroviruses were detected in 7 of 7 and 3 of 8 WWTPs and building-level samples, respectively, with child care center A and nursing home D obtaining the greatest relative abundance among samples.

Astroviruses cause a significant burden mainly among pediatric and elderly populations (Vu et al., 2017). They are a leading cause of gastroenteritis in children (behind rotavirus and norovirus) and could account for 10% of all gastroenteritis outbreaks in child care centers in the U.S. (Lyman et al., 2009). Similarly, elderly populations are at higher risk of gastroenteritis caused by bacterial and viral infections, and individuals in nursing homes face an increased risk of mortality from such infections mainly due to dehydration and comorbidities (Montoya and Mody, 2011).

Further, nursing home E obtained the greatest number of betacoronavirus reads. Severe acute respiratory syndrome coronavirus 2 (SARS-CoV-2), the causative agent of coronavirus disease 2019 (COVID-19), was the most abundant species detected in the betacornavirus genus (data not shown). Since the start of the COVID-19 pandemic, wastewater monitoring of SARS-CoV-2 has played a critical role in early detection of outbreaks and rapid implementation of public health measures (Barrios et al., 2021; Hopkins et al., 2023; Wurtzer et al., 2022).

Interestingly, reads associated with lentivirus were detected in both child care centers and were most abundant in child care center B. The lentivirus genus contains the human immunodeficiency virus (HIV), which is the only species of lentiviruses that affects humans. Targeted metagenomics identified reads homologous to HIV in several wastewater samples including both child care center samples, suggesting the presence of the virus in wastewater and within the community. Perinatal transmission is the most common route among infected children (Deeks et al., 2015). Although newly infected HIV cases in children have declined since its initiation in 1970, HIV remains an ongoing pandemic (Deeks et al., 2015).

Few studies have detected, or focused on methods to detect, HIV in wastewater (Ansari et al., 1992; Kadadou et al., 2022; McCall et al., 2020; Preston et al., 1991). Wastewater surveillance of HIV in high-risk areas could facilitate an increase in status awareness, early application of retroactive and antiviral drugs, and decrease transmission of the viruses in communities. Indeed, it is important to consider the contribution of adults (e.g., teachers, faculty, administrators) when it comes to assessing the prevalence of viruses and age-related disease burdens in school samples, nonetheless wastewater monitoring can be utilized to understand the circulation of clinically important viral pathogens affecting K-12 populations.

Orthohepadnavirus, cytomegalovirus, and, mupapillomavirus were the only genera detected in building-level samples that were undetected in WWTP samples. Cytomegalovirus and mupapillomavirus are associated with retinal lesions in immunocompromised individuals (Dioverti and Razonable, 2016) and skin warts (McBride, 2022) were only detected in nursing home E and the jail samples, respectively. Albeit detected in 4 building-level samples, the jail was the only sample that contained reads homologous to the hepatitis B virus (HBV) in the orthohepadnavirus genus (data not shown). HBV is the causative agent of hepatitis B and can remain in the body for prolonged periods of time. It is primarily transmitted through serum and blood and is the most common chronic viral infection worldwide (Trépo et al., 2014). Incarcerated populations have a disproportionate HBV burden, with an estimated seroprevalence of currently or previously infected inmates of > 40% in the U.S. (Smith et al., 2017). HBV is rarely detected in wastewater samples (Symonds et al., 2009), which suggests that sensitive, targeted metagenomics can be used as a screening tool for both rare and abundant viruses for public health surveillance.

### 3.4. Genetic diversity of astroviruses in wastewater

Near complete and whole-genome contigs were clustered with HAstV-1 and -3 reference sequences (Figure 5). HAstV-1 is one of the most common serotypes isolated from pediatric patients (Cortez et al., 2017; de Grazia et al., 2004; Li et al., 2010) and is frequently detected in wastewater (Hata et al., 2018; Meleg et al., 2006; Ng et al., 2012; Zhou et al., 2014). Although more studies are needed to thoroughly understand differences in virus behaviors between HAstV serotypes, one study found HAstV-3 to be associated with higher fecal concentrations and more persistent and severe gastroenteritis as compared to the other HAstVs (Caballero et al., 2003). Although detected less frequently than HAstV-1, HAstV-3 has also been detected in wastewater samples (Hata et al., 2018). The Child Care Center A-1 contig did not form a cluster with any of the reference sequences. This could be due to further assembly errors or the presence of a divergent sequence. Nonetheless, targeted metagenomics was able to generate putative whole genome contigs at the species level for astroviruses and identify serotypes in wastewater samples.

**Figure 5.**
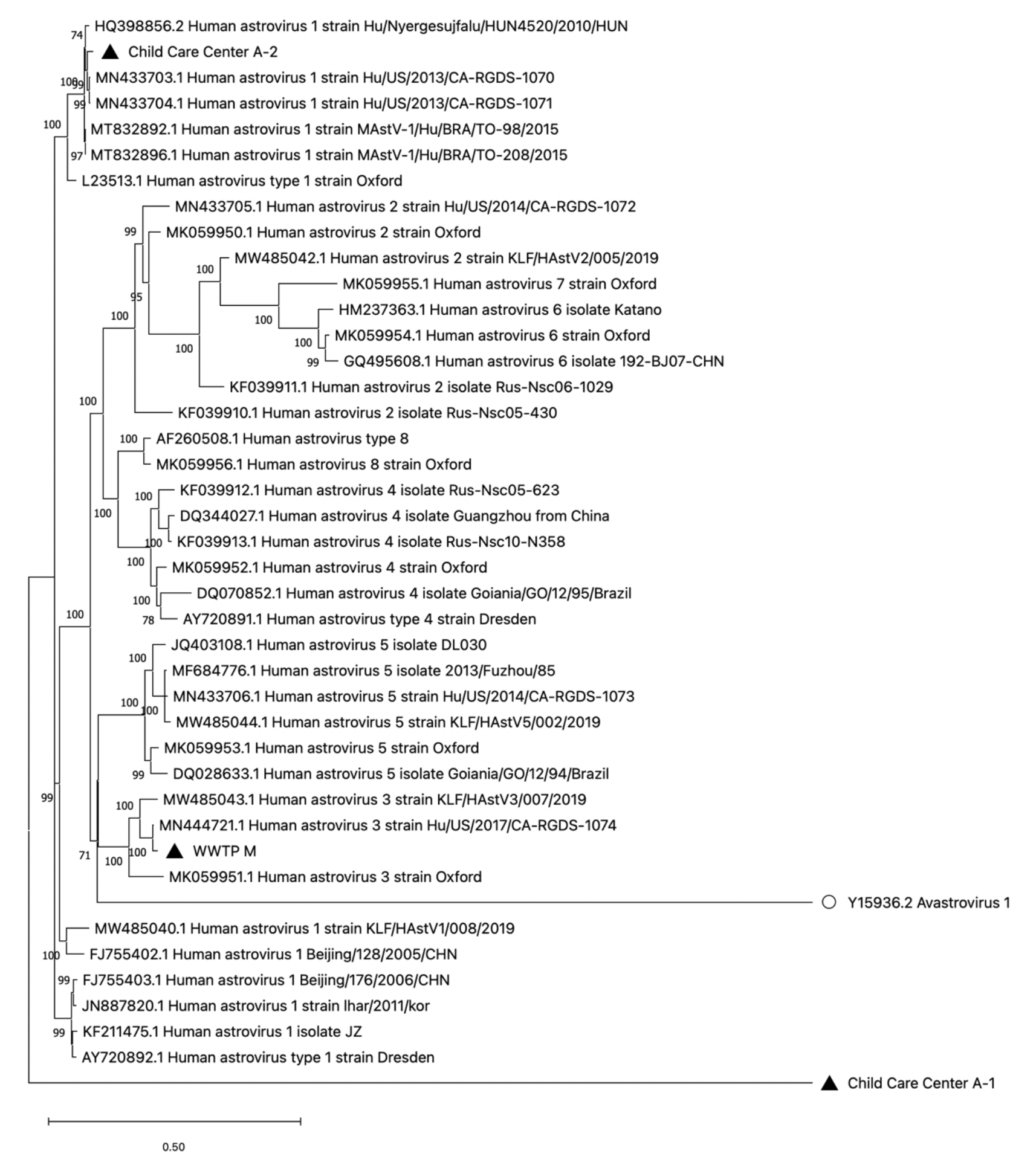
Maximum likelihood phylogenetic tree based on complete genomes of mamastrovirus 1 using the generalized time-reversible model in Fasttree 2.1.11. The tree was constructed with 38 reference sequences from GenBank and 3 putative near complete or complete whole genome contigs from this study. Contigs are denoted with a black triangle. Avastrovirus 1 (white circle) was used as an outgroup. The number on the branches represent the bootstrap percentage and the scale bar refers to the phylogenetic distance in nucleotide substitutions per site. MEGA 11.0 was used for tree visualization only.

### 3.5. Limitations and implications

While we were able to identify important human viruses in building-level samples, more samples of the same community type are needed to statistically evaluate associations between community type and viruses detected. For this reason, determining statistical associations between the viruses detected and demographic groups was outside the scope of this study. However, virus profiles noticeably varied among building-level samples and several viruses detected at the building-level, including HBV, were not detected in WWTP samples. This suggests that building-level sampling can increase the chances of detecting viruses not commonly detected in centralized wastewater samples. Overall, targeted metagenomics increased the number of reads and unique vertebrate viral families in wastewater samples when compared to untargeted metagenomics. Additionally, species-level astrovirus reads produced putative whole-genome contigs with homologies with HAstV-1 and -3 serotypes. This study demonstrates that targeted metagenomic sequencing of WWTP and building-level samples can be used as an initial screening step for public health surveillance and potentially to classify abundant viruses at the subspecies level.

## Data Availability

All data produced in the present study are available upon reasonable request to the authors.

## Acknowledgment

We’d like to thank Houston Public Works and the Houston Health Department for providing wastewater samples. We’d also like to thank Bryce Kille and Advait Balaji in Dr. Todd Treangen’s group for advising on the homology and phylogenetic analyses of astroviruses. Thanks to the Genome Technology Access Center at McDonnell Genome Institute team at Washington University, and specifically Elizabeth Appelbaum and Andrew Emory for providing targeted and untargeted wastewater sequencing data. Lastly, we’d like to thank Madeline Wolken, Lauren Bauhs, Russell Carlson, Kyle Palmer, and Whitney Rich in Lauren Stadler’s lab group at Rice University for their assistance with wastewater sample collection and processing.

## Funding Sources

This work was supported by the Houston Health Department and grants from the National Science Foundation (CBET 2029025) and seed funds from Rice University.

## Supporting Information

Appendix 1: Protocol for Two-Strand cDNA Reverse Transcription for Targeted Metagenomic Sequencing

Appendix 2: Supplementary Table and Figures

Figure S1. Integrative Genomics Viewer (IGV) plot of BWA-MEM read coverage alignment for putative whole genome contig for the Child Care Center A-1 sample.

Figure S2. Integrative Genomics Viewer (IGV) plot of BWA-MEM read coverage alignment for putative whole genome contig for the Child Care Center A-2 sample.

Figure S3. Integrative Genomics Viewer (IGV) plot of BWA-MEM read coverage alignment for putative whole genome contig for the WWTP M sample.

Figure S4. Number of unique vertebrate families detected between electronegative membrane filtered (MF), directly extracted (DE), targeted with probe-based capture, and untargeted samples.

Figure S5. Percent of vertebrate reads detected between electronegative membrane filtered (MF), directly extracted (DE), targeted with probe-based capture, and untargeted samples.

Table S1. Summary of contig coverage results from alignment with BWA-MEM and visualization with Integrative Genomics Viewer (IGV).

Table S2. Summary of read count per vertebrate virus family in untargeted/electronegative membrane filtered (MF) and targeted/MF samples for WWTPs M and O.

Table S3. Summary of read count per vertebrate virus family in targeted/directly extracted (DE) and targeted/electronegative membrane filtered (MF) samples for WWTPs M and O.

Table S4. Total number of sequencing reads per sample assigned to viruses using the ViroMatch pipeline.

Table S5. Summary human viruses detected in building-level and WWTP samples and their associated route of transmission.

## Notes

### Competing Interest Statement

The authors have declared no competing interest.

## References

Ansari, S.A., Farrah, S.R., Chaudhry, G.R., 1992. Presence of Human Immunodeficiency Virus Nucleic Acids in Wastewater and Their Detection by Polymerase Chain Reaction. Appl Environ Microbiol 58, 3984–3990.

Aw, T.G., Howe, A., Rose, J.B., 2014. Metagenomic approaches for direct and cell culture evaluation of the virological quality of wastewater. J Virol Methods 210, 15–21. https://doi.org/10.1016/j.jviromet.2014.09.017

Barrios, R.E., Lim, C., Kelley, M.S., Li, X., 2021. SARS-CoV-2 concentrations in a wastewater collection system indicated potential COVID-19 hotspots at the zip code level. Science of the Total Environment 800. https://doi.org/10.1016/j.scitotenv.2021.149480

Bibby, K., Peccia, J., 2013. Identification of viral pathogen diversity in sewage sludge by metagenome analysis. Environ Sci Technol 47, 1945–1951. https://doi.org/10.1021/es305181x

Bisseux, M., Colombet, J., Mirand, A., Roque-Afonso, A.M., Abravanel, F., Izopet, J., Archimbaud, C., Peigue-Lafeuille, H., Debroas, D., Bailly, J.L., Henquell, C., 2018. Monitoring human enteric viruses in wastewater and relevance to infections encountered in the clinical setting: A one-year experiment in central France, 2014 to 2015. Eurosurveillance 23, 1–11. https://doi.org/10.2807/1560-7917.ES.2018.23.7.17-00237

Caballero, S., Guix, S., El-Senousy, W.M., Calicó, I., Pintó, R.M., Bosch, A., 2003. Persistent gastroenteritis in children infected with astrovirus: Association with serotype-3 strains. J Med Virol 71, 245–250. https://doi.org/10.1002/jmv.10476

Cantalupo, P.G., Calgua, B., Zhao, G., Hundesa, A., Wier, A.D., Katz, J.P., Grabe, M., Hendrix, R.W., Girones, R., Wang, D., Pipas, J.M., 2011. Raw sewage harbors diverse viral populations. mBio 2, 1–11. https://doi.org/10.1128/mBio.00180-11

Choi, P.M., Tscharke, B., Samanipour, S., Hall, W.D., Gartner, C.E., Mueller, J.F., Thomas, K. v., O’Brien, J.W., 2019. Social, demographic, and economic correlates of food and chemical consumption measured by wastewater-based epidemiology. Proc Natl Acad Sci U S A 116, 21864–21873. https://doi.org/10.1073/pnas.1910242116

Cortez, V., Freiden, P., Gu, Z., Adderson, E., Hayden, R., Schultz-Cherry, S., 2017. Persistent infections with diverse co-circulating astroviruses in pediatric oncology patients, Memphis, Tennessee, USA. Emerg Infect Dis 23, 288–290. https://doi.org/10.3201/eid2302.161436

de Grazia, S., Giammanco, G.M., Colomba, C., Cascio, A., Arista, S., 2004. Molecular epidemiology of astrovirus infection in Italian children with gastroenteritis. Clinical Microbiology and Infection 10, 1025–1029. https://doi.org/10.1111/j.1469-691.2004.00995.x

Deeks, S.G., Overbaugh, J., Phillips, A., Buchbinder, S., 2015. HIV infection. Nat Rev Dis Primers 1. https://doi.org/10.1038/nrdp.2015.35

Dioverti, M.V., Razonable, R.R., 2016. Cytomegalovirus. Microbiol Spectr 4. https://doi.org/10.1128/microbiolspec.DMIH2-0022-2015

Donato, C., Vijaykrishna, D., 2017. The broad host range and genetic diversity of mammalian and avian astroviruses. Viruses. https://doi.org/10.3390/v9050102

Farkas, K., Cooper, D.M., McDonald, J.E., Malham, S.K., de Rougemont, A., Jones, D.L., 2018. Seasonal and spatial dynamics of enteric viruses in wastewater and in riverine and estuarine receiving waters. Science of the Total Environment 634, 1174–1183. https://doi.org/10.1016/j.scitotenv.2018.04.038

Fernandez-Cassi, X., Martínez-Puchol, S., Silva-Sales, M., Cornejo, T., Bartolome, R., Bofill-Mas, S., Girones, R., 2020. Unveiling viruses associated with gastroenteritis using a metagenomics approach. Viruses 12. https://doi.org/10.3390/v12121432

Fernandez-Cassi, X., Scheidegger, A., Bänziger, C., Cariti, F., Tuñas Corzon, A., Ganesanandamoorthy, P., Lemaitre, J.C., Ort, C., Julian, T.R., Kohn, T., 2021. Wastewater monitoring outperforms case numbers as a tool to track COVID-19 incidence dynamics when test positivity rates are high. Water Res 200. https://doi.org/10.1016/j.watres.2021.117252

Galani, A., Aalizadeh, R., Kostakis, M., Markou, A., Alygizakis, N., Lytras, T., Adamopoulos, P.G., Peccia, J., Thompson, D.C., Kontou, A., Karagiannidis, A., Lianidou, E.S., Avgeris, M., Paraskevis, D., Tsiodras, S., Scorilas, A., Vasiliou, V., Dimopoulos, M.A., Thomaidis, N.S., 2022. SARS-CoV-2 wastewater surveillance data can predict hospitalizations and ICU admissions. Science of the Total Environment 804. https://doi.org/10.1016/j.scitotenv.2021.150151

Girón-Guzmán, I., Díaz-Reolid, A., Cuevas-Ferrando, E., Falcó, I., Cano-Jiménez, P., Comas, I., Pérez-Cataluña, A., Sánchez, G., 2023. Evaluation of two different concentration methods for surveillance of human viruses in sewage and their effects on SARS-CoV-2 sequencing. Science of the Total Environment 862. https://doi.org/10.1016/j.scitotenv.2022.160914

Hall, R.J., Wang, J., Todd, A.K., Bissielo, A.B., Yen, S., Strydom, H., Moore, N.E., Ren, X., Huang, Q.S., Carter, P.E., Peacey, M., 2014. Evaluation of rapid and simple techniques for the enrichment of viruses prior to metagenomic virus discovery. J Virol Methods 195, 194–204. https://doi.org/10.1016/j.jviromet.2013.08.035

Haramoto, E., Kitajima, M., Hata, A., Torrey, J.R., Masago, Y., Sano, D., Katayama, H., 2018. A review on recent progress in the detection methods and prevalence of human enteric viruses in water. Water Res 135, 168–186. https://doi.org/10.1016/j.watres.2018.02.004

Hata, A., Kitajima, M., Haramoto, E., Lee, S., Ihara, M., Gerba, C.P., Tanaka, H., 2018. Next-generation amplicon sequencing identifies genetically diverse human astroviruses, including recombinant strains, in environmental waters. Sci Rep 8. https://doi.org/10.1038/s41598-018-30217-y

Hellmér, M., Paxéus, N., Magnius, L., Enache, L., Arnholm, B., Johansson, A., Bergström, T., Norder, H., 2014. Detection of pathogenic viruses in sewage provided early warnings of hepatitis A virus and norovirus outbreaks. Appl Environ Microbiol 80, 6771–6781. https://doi.org/10.1128/AEM.01981-14

Hjelmsø, M.H., Hellmér, M., Fernandez-Cassi, X., Timoneda, N., Lukjancenko, O., Seidel, M., Elsässer, D., Aarestrup, F.M., Löfström, C., Bofill-Mas, S., Abril, J.F., Girones, R., Schultz, A.C., 2017. Evaluation of methods for the concentration and extraction of viruses from sewage in the context of metagenomic sequencing. PLoS One 12, 1–17. https://doi.org/10.1371/journal.pone.0170199

Hopkins, L., Persse, D., Caton, K., Ensor, K., Schneider, R., McCall, C., Stadler, L.B., 2023. Citywide wastewater SARS-CoV-2 levels strongly correlated with multiple disease surveillance indicators and outcomes over three COVID-19 waves. Science of the Total Environment 855. https://doi.org/10.1016/j.scitotenv.2022.158967

Kadadou, D., Tizani, L., Wadi, V.S., Banat, F., Alsafar, H., Yousef, A.F., Barceló, D., Hasan, S.W., 2022. Recent advances in the biosensors application for the detection of bacteria and viruses in wastewater. J Environ Chem Eng 10. https://doi.org/10.1016/j.jece.2021.107070

Kirby, A.E., Walters, M.S., Jennings, W.C., Fugitt, R., LaCross, N., Mattioli, M., Marsh, Z.A., Roberts, V.A., Mercante, J.W., Yoder, J., Hill, V.R., 2021. Using Wastewater Surveillance Data to Support the COVID-19 Response — United States, 2020–2021. Morbidity and Mortality Weekly Report 70, 1242–1244.

LaTurner, Z.W., Zong, D.M., Kalvapalle, P., Gamas, K.R., Terwilliger, A., Crosby, T., Ali, P., Avadhanula, V., Santos, H.H., Weesner, K., Hopkins, L., Piedra, P.A., Maresso, A.W., Stadler, L.B., 2021. Evaluating recovery, cost, and throughput of different concentration methods for SARS-CoV-2 wastewater-based epidemiology. Water Res 197. https://doi.org/10.1016/j.watres.2021.117043

Li, C.Y., Liu, N., Guo, W.D., Yu, Q., Wang, W.R., Song, Z.Z., Yan, H., Luo, Y., Lu, A.T., Li, H.Y., Zhu, L., Duan, Z.J., 2010. Outbreak of neonatal gastroenteritis associated with astrovirus serotype 1 at a hospital in inner Mongolia, China. J Clin Microbiol 48, 4306–4309. https://doi.org/10.1128/JCM.00797-10

Lyman, W.H., Walsh, J.F., Kotch, J.B., Weber, D.J., Gunn, E., Vinjé, J., 2009. Prospective Study of Etiologic Agents of Acute Gastroenteritis Outbreaks in Child Care Centers. Journal of Pediatrics 154, 253–257. https://doi.org/10.1016/j.jpeds.2008.07.057

Martínez-Puchol, S., Rusiñol, M., Fernández-Cassi, X., Timoneda, N., Itarte, M., Andrés, C., Antón, A., Abril, J.F., Girones, R., Bofill-Mas, S., 2020. Characterisation of the sewage virome: comparison of NGS tools and occurrence of significant pathogens. Science of the Total Environment 713. https://doi.org/10.1016/j.scitotenv.2020.136604

Matus, M., Duvallet, C., Soule, M.K., Kearney, S.M., Endo, N., Ghaeli, N., Brito, I., Ratti, C., Kujawinski, E.B., Alm, E.J., 2019. 24-hour multi-omics analysis of residential sewage reflects human activity and informs public health. bioRxiv. https://doi.org/10.1101/728022

McBride, A.A., 2022. Human papillomaviruses: diversity, infection and host interactions. Nat Rev Microbiol 20, 95–108. https://doi.org/10.1038/s41579-021-00617-5

McCall, C., Wu, H., Miyani, B., Xagoraraki, I., 2020. Identification of multiple potential viral diseases in a large urban center using wastewater surveillance. Water Res 184. https://doi.org/10.1016/j.watres.2020.116160

McCall, C., Wu, H., O’Brien, E., Xagoraraki, I., 2021. Assessment of enteric viruses during a hepatitis outbreak in Detroit MI using wastewater surveillance and metagenomic analysis. J Appl Microbiol 131, 1539–1554. https://doi.org/10.1111/jam.15027

Meleg, E., Jakab, F., Kocsis, B., Bányai, K., Melegh, B., Szucs, G., 2006. Human astroviruses in raw sewage samples in Hungary. J Appl Microbiol. https://doi.org/10.1111/j.1365-2672.2006.02997.x

Mondal, S., Feirer, N., Brockman, M., Preston, M.A., Teter, S.J., Ma, D., Goueli, S.A., Moorji, S., Saul, B., Cali, J.J., 2021. A direct capture method for purification and detection of viral nucleic acid enables epidemiological surveillance of SARS-CoV-2. Science of the Total Environment 795. https://doi.org/10.1016/j.scitotenv.2021.148834

Montoya, A., Mody, L., 2011. Common infections in nursing homes: A review of current issues and challenges. Aging health 7, 889–899. https://doi.org/10.2217/ahe.11.80

Newton, R.J., McLellan, S.L., Dila, D.K., Vineis, J.H., Morrison, H.G., Murat Eren, A., Sogin, M.L., 2015. Sewage reflects the microbiomes of human populations. mBio 6. https://doi.org/10.1128/mBio.02574-14

Ng, T.F.F., Marine, R., Wang, C., Simmonds, P., Kapusinszky, B., Bodhidatta, L., Oderinde, B.S., Wommack, K.E., Delwart, E., 2012. High variety of known and new RNA and DNA viruses of diverse origins in untreated sewage. J Virol 86, 12161–12175. https://doi.org/10.1128/jvi.00869-12

O’Brien, E., Nakyazze, J., Wu, H., Kiwanuka, N., Cunningham, W., Kaneene, J.B., Xagoraraki, I., 2017. Viral diversity and abundance in polluted waters in Kampala, Uganda. Water Res 127, 41–49. https://doi.org/10.1016/j.watres.2017.09.063

Paskey, A.C., Frey, K.G., Schroth, G., Gross, S., Hamilton, T., Bishop-Lilly, K.A., 2019. Enrichment post-library preparation enhances the sensitivity of high-throughput sequencing-based detection and characterization of viruses from complex samples. BMC Genomics 20. https://doi.org/10.1186/s12864-019-5543-2

Peccia, J., Zulli, A., Brackney, D.E., Grubaugh, N.D., Kaplan, E.H., Casanovas-Massana, A., Ko, A.I., Malik, A.A., Wang, D., Wang, M., Warren, J.L., Weinberger, D.M., Arnold, W., Omer, S.B., 2020. Measurement of SARS-CoV-2 RNA in wastewater tracks community infection dynamics. Nat Biotechnol 38, 1164–1167. https://doi.org/10.1038/s41587-020-0684-z

Preston, D.R., Farrah, S.R., Bitton, G., Chaudhry, G. Rasul., 1991. Detection of nucleic acids homologous to human immunodefkiency virus in wastewater”. J Virol Methods 33, 383–390.

Prevost, B., Lucas, F.S., Goncalves, A., Richard, F., Moulin, L., Wurtzer, S., 2015. Large scale survey of enteric viruses in river and waste water underlines the health status of the local population. Environ Int 79, 42–50. https://doi.org/10.1016/j.envint.2015.03.004

R Core Team, 2022. R: A language and environment for statistical computing. Vienna, Austria.

Segelhurst, E., Bard, J.E., Pillsbury, A.N., Mahbubul Alam, M., Lamb, A., Zhu, C., Pohlman, A., Boccolucci, A., Emerson, J., Yergeau, D.A., Nowak, N.J., Bradley, I.M., Surtees, J.A., Ye, Y., 2022. Improved Robustness of SARS-CoV-2 Whole-Genome Sequencing from Wastewater with a Nonselective Virus Concentration Method. medRxiv. https://doi.org/10.1101/2022.09.07.22279692

Smith, J.M., Uvin, A.Z., Macmadu, A., Rich, J.D., 2017. Epidemiology and Treatment of Hepatitis B in Prisoners. Curr Hepatol Rep 16, 178–183. https://doi.org/10.1007/s11901-017-0364-8

Spurbeck, R.R., Minard-Smith, A., Catlin, L., 2021. Feasibility of neighborhood and building scale wastewater-based genomic epidemiology for pathogen surveillance. Science of the Total Environment 789. https://doi.org/10.1016/j.scitotenv.2021.147829

Sutton, T.D.S., Clooney, A.G., Ryan, F.J., Ross, R.P., Hill, C., 2019. Choice of assembly software has a critical impact on virome characterisation. Microbiome 7. https://doi.org/10.1186/s40168-019-0626-5

Symonds, E.M., Griffin, D.W., Breitbart, M., 2009. Eukaryotic viruses in wastewater samples from the United States. Appl Environ Microbiol 75, 1402–1409. https://doi.org/10.1128/AEM.01899-08

Tao, Z., Lin, X., Liu, Y., Ji, F., Wang, S., Xiong, P., Zhang, L., Xu, Q., Xu, A., Cui, N., 2022. Detection of multiple human astroviruses in sewage by next generation sequencing. Water Res 218. https://doi.org/10.1016/j.watres.2022.118523

Trépo, C., Chan, H.L.Y., Lok, A., 2014. Hepatitis B virus infection. The Lancet 384, 2053–2063. https://doi.org/10.1016/S0140-6736(14)60220-8

Vu, D.L., Bosch, A., Pintó, R.M., Guix, S., 2017. Epidemiology of classic and novel human astrovirus: Gastroenteritis and beyond. Viruses 9. https://doi.org/10.3390/v9020033

Wang, D., Urisman, A., Liu, Y.T., Springer, M., Ksiazek, T.G., Erdman, D.D., Mardis, E.R., Hickenbotham, M., Magrini, V., Eldred, J., Latreille, J.P., Wilson, R.K., Ganem, D., DeRisi, J.L., 2003. Viral discovery and sequence recovery using DNA microarrays. PLoS Biol 1. https://doi.org/10.1371/journal.pbio.0000002

Westerhoff, P., Lee, S., Yang, Y., Gordon, G.W., Hristovski, K., Halden, R.U., Herckes, P., 2015. Characterization, Recovery Opportunities, and Valuation of Metals in Municipal Sludges from U.S. Wastewater Treatment Plants Nationwide. Environ Sci Technol 49, 9479–9488. https://doi.org/10.1021/es505329q

Wurtzer, S., Waldman, P., Levert, M., Cluzel, N., Almayrac, J.L., Charpentier, C., Masnada, S., Gillon-Ritz, M., Mouchel, J.M., Maday, Y., Boni, M., Moulin, L., Marechal, V., le Guyader, S., Bertrand, I., Gantzer, C., Descamps, D., Charpentier, Charlotte, Houhou, N., Visseaux, B., Calvez, V., Marcelin, A.G., Marot, S., Jary, A., Chaix, M.L., Delaugerre, C., Salmona, M., Legoff, J., Gault, E., Rameix-Welti, M.A., L’honneur, A.S., Mariaggi, A.A., Rozenberg, F., Roque, A.M., Mouna, L., Fenaux, H., Leruez-Ville, M., Morand-Joubert, L., Lambert-Niclot, S., Schnuriger, A., Dubois, C., Bourgeois-Nicolaos, N., Brichler, S., Delagrèverie, H., Bouvier-Alias, M., Fourati, S., Rodriguez, C., Pawlotsky, J.M., 2022. SARS-CoV-2 genome quantification in wastewaters at regional and city scale allows precise monitoring of the whole outbreaks dynamics and variants spreading in the population. Science of the Total Environment 810. https://doi.org/10.1016/j.scitotenv.2021.152213

Wylie, K.M., Wylie, T.N., Buller, R., Herter, B., Cannella, M.T., Storcha, G.A., 2018. Detection of Viruses in Clinical Samples by Use of Metagenomic Sequencing and Targeted Sequence Capture. J Clin Microbiol 56. https://doi.org/10.1128/JCM.01123-18

Wylie, T.N., Wylie, K.M., 2021. ViroMatch: A Computational Pipeline for the Detection of Viral Sequences from Complex Metagenomic Data. Microbiol Resour Announc 10. https://doi.org/10.1128/mra.01468-20

Wylie, T.N., Wylie, K.M., Herter, B.N., Storch, G.A., 2015. Enhanced virome sequencing using targeted sequence capture. Genome Res 25, 1910–1920. https://doi.org/10.1101/gr.191049.115

Xu, B., Liu, L., Huang, X., Ma, H., Zhang, Y., Du, Y., Wang, P., Tang, X., Wang, H., Kang, K., Zhang, S., Zhao, G., Wu, W., Yang, Y., Chen, H., Mu, F., Chen, W., 2011. Metagenomic analysis of fever, thrombocytopenia and leukopenia syndrome (FTLS) in Henan province, China: Discovery of a new bunyavirus. PLoS Pathog 7. https://doi.org/10.1371/journal.ppat.1002369

Zhao, L., Zou, Y., Li, Y., Miyani, B., Spooner, M., Gentry, Z., Jacobi, S., David, R.E., Withington, S., McFarlane, S., Faust, R., Sheets, J., Kaye, A., Broz, J., Gosine, A., Mobley, P., Busch, A.W.U., Norton, J., Xagoraraki, I., 2022. Five-week warning of COVID-19 peaks prior to the Omicron surge in Detroit, Michigan using wastewater surveillance. Science of the Total Environment 844. https://doi.org/10.1016/j.scitotenv.2022.157040

Zhou, N., Lin, X., Wang, S., Wang, H., Li, W., Tao, Z., Xu, A., 2014. Environmental surveillance for human astrovirus in Shandong Province, China in 2013. Sci Rep 4. https://doi.org/10.1038/srep07539

