## Supplemental Information for "Targeted metagenomic sequencing for detection of vertebrate viruses in wastewater for public health surveillance"

Mailing address:

6100 Main Street, MS-519

Houston, TX 77005

Supporting Information
Pages: 14
Figures: 5
Tables: 5

### Supporting Information

#### Appendix 1: Protocol for Two-Strand cDNA Reverse Transcription for Targeted Metagenomic Sequencing

This protocol was taken, in part, from Wang et al., 2003 with modifications to the random primer used and the thermocycling conditions for first-strand synthesis.

#### Reagents

- ☐ Reverse transcriptase (SuperScript III, Invitrogen Cat No. 18080-044 for 50 rxn)
- ☐ 10 mM dNTP mix (Promega)
- ☐ DEPC-treated water (Invitrogen Cat No. 10813-012)
- ☐ 40 pmol/µl Primer A: NNN NNN NN
- ☐ Sequenase (Affymetrix Cat No. 70775Y 200 UN)
- ☐ RNaseOUT Recombinant Ribonuclease Inhibitor (Invitrogen Cat No. 10777-019)

#### Protocol

*First-strand synthesis with RT*

1. Add 40 µM of Primer A to 5µl of extracted nucleic acid and DEPC-treated water to a final volume of 10µl.
2. Heat samples to 65° C for 5 min in a thermocycler
3. Let cool at room temperature for 5 min.

4. Add 10µl of the following master mix to each sample:
   a. 4µl 5X RT buffer
   b. 1µl 10 mM dNTP
   c. 0.5µl water
   d. 1.5µl 0.1M DTT
   e. 2µl SSIII RT
   f. 1µl RNaseOUT

5. Incubate at 42° C for 60 min. Heat sample to 94° C for 2 min (to abort the RT reaction). Rapidly cool to 10° C and hold at 10° C for 5 min.
6. Keep samples on ice to maintain a temperature of ~10° C.

*Second-strand synthesis with Sequenase*

1. Create the following mixtures:

<u>Sequenase mix</u>

   a. 2µl 5X Sequenase buffer

b. 7.7µl water

c. 0.3µl Sequenase

Diluted sequenase mix

Make diluted Sequenase mix with a 1:4 dilution using Sequenase dilution buffer and Sequenase. Prepare enough to supply all samples with 1.2 µl of diluted mix.

2. Add 10µl Sequenase mix for a total reaction volume of 30µl:
    a. 2µl 5X Sequenase buffer
    b. 7.7µl water
    c. 0.3µl Sequenase

3. Run samples with the following conditions in a thermocycler:
    a. Ramp from 10°C to 37°C over 8 min.
    b. Hold at 37°C for 8 min, rapid ramp to 94°C and hold for 2 min.
    **c.** Rapid ramp to 10°C and hold for 5 min at 10°C while adding 1.2µl of diluted Sequenase (1:4 dilution). Open the thermocycler lid while on the "hold for 5 min at 10ºC" step and add diluted Sequenase to each tube. Close lid when finished.
    d. Ramp from 10°C to 37°C over 8 min.
    e. Hold at 37°C for 8 min, ramp to 94°C and hold for 8 min (to inactivate Sequenase). Cool to 10°C (hold).

4. Run Qubit on double-stranded cDNA samples using the appropriate kit. Record concentrations.

**Appendix 2: Supplementary Table and Figures**

Table S1. Summary of contig coverage results from alignment with BWA-MEM and visualization with Integrative Genomics Viewer (IGV).

| Sample | Contig Length (bp) | Amended Contig Length (bp) | GC-content (%) | Average Read Depth | Standard Deviation of Read Depth | Contig Coverage (%) |
|---|---|---|---|---|---|---|
| Child Care Center A-1 | 6692 | | 43.28 | 60.25 | 49.39 | 100 |
| Child Care Center A-2 | 6351 | 5833 | 44.99 | 981.57 | 897.22 | 100 |
| WWTP M | 6863 | 4436 | 43.80 | 477.83 | 458.32 | 99.08 |

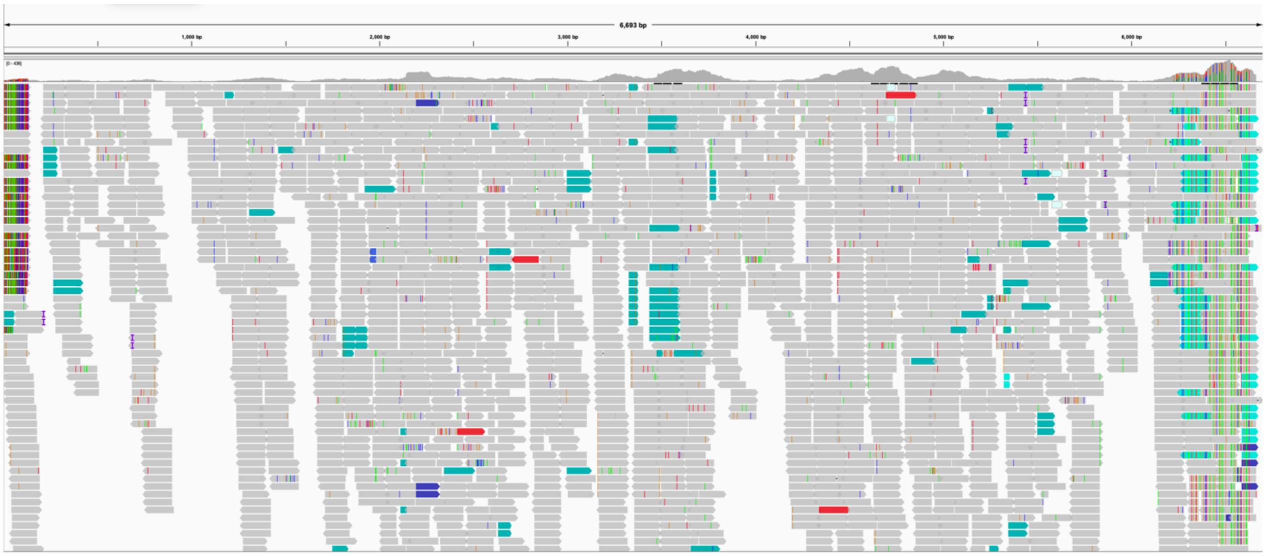

Figure S1. Integrative Genomics Viewer (IGV) plot of BWA-MEM read coverage alignment for putative whole genome contig for the Child Care Center A-1 sample.

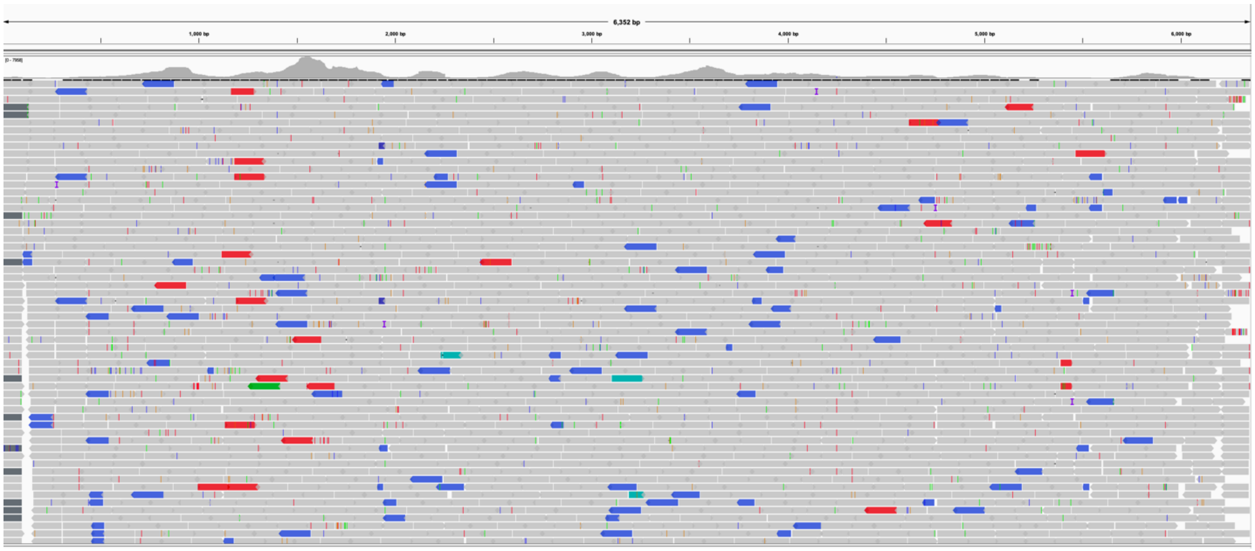

Figure S2. Integrative Genomics Viewer (IGV) plot of BWA-MEM read coverage alignment for putative whole genome contig for the Child Care Center A-2 sample.

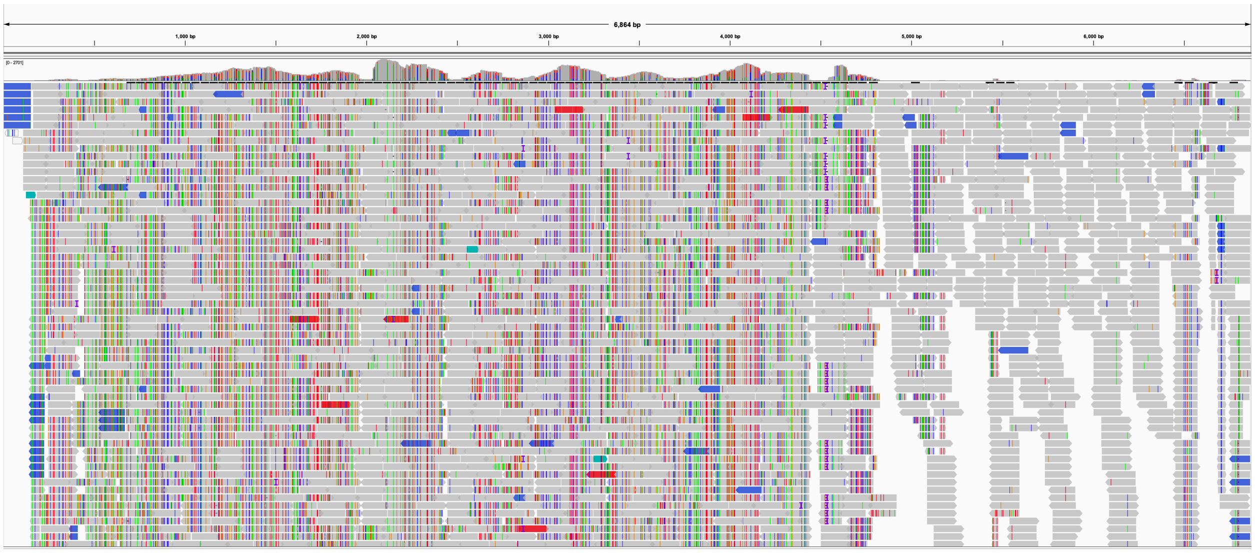

Figure S3. Integrative Genomics Viewer (IGV) plot of BWA-MEM read coverage alignment for putative whole genome contig for the WWTP M sample.

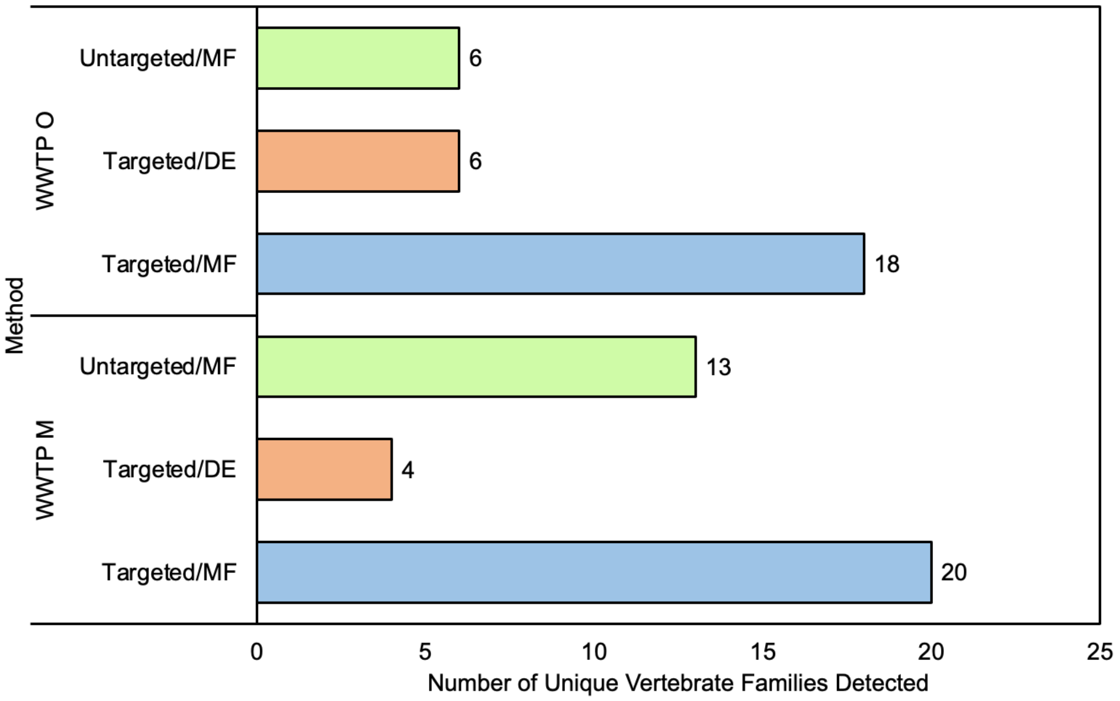


Figure S4. Number of unique vertebrate families detected between electronegative membrane filtered (MF), directly extracted (DE), targeted with probe-based capture, and untargeted samples.

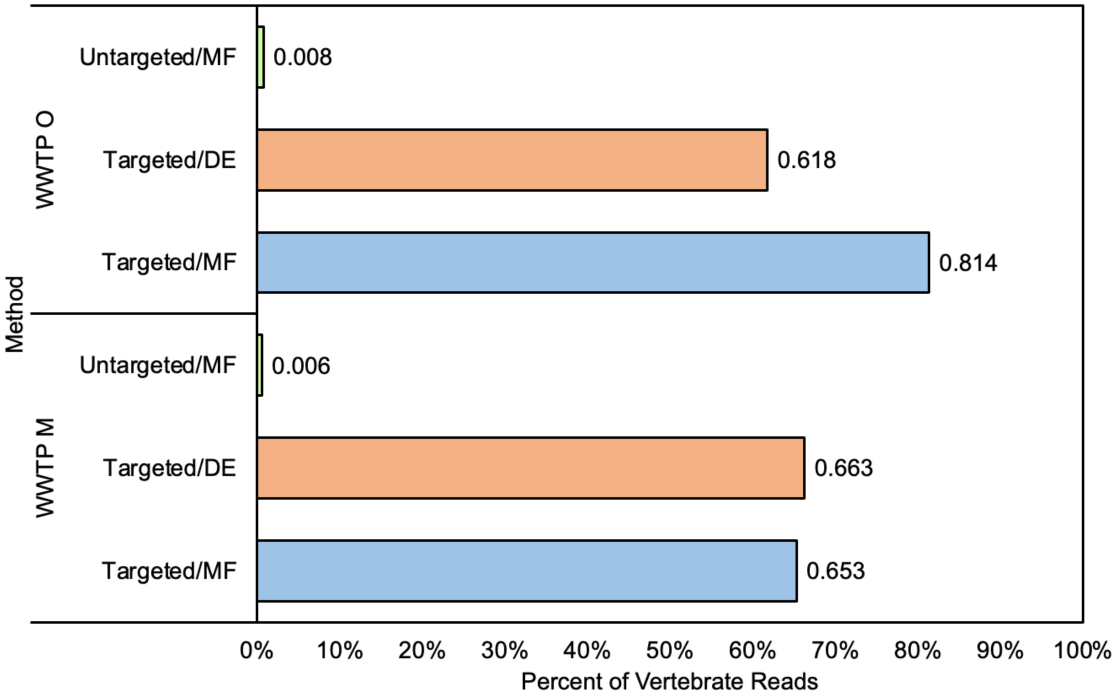

Figure S5. Percent of vertebrate reads detected between electronegative membrane filtered (MF), directly extracted (DE), targeted with probe-based capture, and untargeted samples.

Table S2. Summary of read count per vertebrate virus family in untargeted/electronegative membrane filtered (MF) and targeted/MF samples for WWTPs M and O.

| | WWTP M | | WWTP O | |
|---|---|---|---|---|
| **Viral families** | **Untargeted** | **Targeted** | **Untargeted** | **Targeted** |
| Adenoviridae | 4 | 16309 | 1 | 36383 |
| Anelloviridae | 0 | 151 | 0 | 135 |
| Asfarviridae | 25 | 3 | 0 | 5 |
| Astroviridae | 5 | 69944 | 0 | 34454 |
| Caliciviridae | 4 | 2019 | 0 | 2299 |
| Circoviridae | 79 | 194 | 32 | 138 |
| Coronaviridae | 0 | 1581 | 0 | 1532 |
| Herpesviridae | 5 | 163 | 0 | 172 |
| Papillomaviridae | 2 | 121 | 0 | 72 |
| Paramyxoviridae | 0 | 4 | 0 | 0 |
| Parvoviridae | 60 | 4916 | 6 | 6745 |
| Picobirnaviridae | 38 | 150 | 30 | 68 |
| Picornaviridae | 2 | 3945 | 2 | 3258 |
| Pneumoviridae | 0 | 2 | 0 | 6 |
| Polyomaviridae | 0 | 598 | 0 | 212 |
| Poxviridae | 319 | 113 | 176 | 32 |
| Reoviridae | 0 | 1786 | 0 | 291 |

| | | | | |
|---|---|---|---|---|
| Retroviridae | 2 | 1 | 0 | 0 |
| Smacoviridae | 2 | 145 | 0 | 171 |
| Tobaniviridae | 0 | 102 | 0 | 58 |

Table S3. Summary of read count per vertebrate virus family in targeted/directly extracted (DE) and targeted/electronegative membrane filtered (MF) samples for WWTPs M and O.

| **Viral families** | **WWTP M** | | **WWTP O** | |
|---|---|---|---|---|
| | **DE** | **MF** | **DE** | **MF** |
| Adenoviridae | 0 | 16309 | 748 | 36383 |
| Anelloviridae | 0 | 151 | 0 | 135 |
| Asfarviridae | 0 | 3 | 0 | 5 |
| Astroviridae | 0 | 69944 | 0 | 34454 |
| Caliciviridae | 0 | 2019 | 30 | 2299 |
| Circoviridae | 0 | 194 | 0 | 138 |
| Coronaviridae | 131 | 1581 | 0 | 1532 |
| Herpesviridae | 7 | 163 | 0 | 172 |
| Papillomaviridae | 10 | 121 | 0 | 72 |
| Paramyxoviridae | 0 | 4 | 0 | 0 |
| Parvoviridae | 0 | 4916 | 4 | 6745 |
| Picobirnaviridae | 0 | 150 | 0 | 68 |
| Picornaviridae | 0 | 3945 | 145 | 3258 |
| Pneumoviridae | 0 | 2 | 0 | 6 |
| Polyomaviridae | 0 | 598 | 23 | 212 |
| Poxviridae | 0 | 113 | 3 | 32 |

| | | | | |
|---|---|---|---|---|
| Reoviridae | 0 | 1786 | 0 | 291 |
| Retroviridae | 11 | 1 | 0 | 0 |
| Smacoviridae | 0 | 145 | 0 | 171 |
| Tobaniviridae | 0 | 102 | 0 | 58 |

Table S4. Total number of sequencing reads per sample assigned to viruses using the ViroMatch pipeline.

| Sample Type | Sample | Total Number of Viral-assigned Reads |
|---|---|---|
| Child Care Center | A | 90734 |
| | B | 6840 |
| High School | C | 27140 |
| Nursing Home | D | 68078 |
| | E | 2379 |
| Homeless Shelter | F | 15206 |
| | G | 15686 |
| Jail | H | 54184 |
| WWTP | I | 12216 |
| | J | 39391 |
| | K | 16651 |
| | L | 101358 |
| | M | 156567 |
| | N | 66422 |
| | O | 105885 |

Table S5. Summary human viruses detected in building-level and WWTP samples and their associated route of transmission.

| Family | Genus | Virus Type (by Primary Route of Transmission in Humans) | Detection | | Ref |
|---|---|---|---|---|---|
| | | | Building-level | WWTP | |

| | | | | | |
|---|---|---|---|---|---|
| Adenoviridae | Mastadenovirus | Enteric, Respiratory | + | + | (Ghebremedhin, 2014) |
| | Unclassified Adenoviridae | Unclassified | + | + | - |
| Anelloviridae | Gyrovirus | Unknown | + | + | (Kaczorowska and van der Hoek, 2020) |
| | Unclassified Anelloviridae | Unclassified | + | + | - |
| | Betatorquevirus | Unknown | + | + | (Kaczorowska and van der Hoek, 2020) |
| | Alphatorquevirus | Unknown | - | + | |
| | Gammatorquevirus | Unknown | - | + | |
| Astroviridae | Mamastrovirus | Enteric | + | + | (Bosch et al., 2014) |
| | Unclassified Astroviridae | Unclassified | + | + | - |
| Caliciviridae | Norovirus | Enteric | + | + | (Glass et al., 2009) |
| | Sapovirus | Enteric | + | + | (Oka et al., 2015) |
| Circoviridae | Cyclovirus | Enteric | + | + | (Li et al., 2010) |
| Coronaviridae | Alphacoronavirus | Respiratory | - | + | (Kutter et al., 2018) |
| | Betacoronavirus | Respiratory | + | + | |
| Hepadnaviridae | Orthohepadnavirus | Bloodborne | + | - | (Smith et al., 2017) |
| Herpesviridae | Roseolovirus | Respiratory, Other | + | + | (de Bolle et al., 2005) |
| | Lymphocryptovirus | Bloodborne, Other | + | + | (Tsao et al., 2015) |
| | Simplexvirus | Other | + | + | (Widener and Whitley, 2014) |
| | Cytomegalovirus | Other | + | - | (Dioverti and Razonable, 2016) |
| | Rhadinovirus | Other | - | + | (Mesri et al., 2010) |

| | | | | | |
|---|---|---|---|---|---|
| Papillomaviridae | Alphapapillomavirus | Other | + | + | (Doorbar et al., 2015) |
| | Betapapillomavirus | Other | + | + | |
| | Gammapapillomavirus | Other | + | + | |
| | Mupapillomavirus | Other | + | - | (Doorbar et al., 2015; McBride, 2022) |
| | Unclassified Papillomaviridae | Unclassified | + | + | - |
| Paramyxoviridae | Respirovirus | Respiratory | - | + | (Rima et al., 2019) |
| | Rubulavirus | Respiratory | - | + | |
| Parvoviridae | Bocaparvovirus | Enteric, Respiratory | + | + | (Qiu et al., 2017) |
| | Dependoparvovirus | Other | - | + | (Jager et al., 2021) |
| | Unclassified Parvovirinae | Unclassified | + | + | - |
| Picobirnaviridae | Picobirnavirus | Enteric | + | + | (Ganesh et al., 2012) |
| Picornaviridae | kobuvirus | Enteric | + | + | (Khamrin et al., 2014) |
| | Parechovirus | Enteric, Respiratory | + | + | (de Crom et al., 2016) |
| | Hepatovirus | Enteric | + | + | (Lemon et al., 2018) |
| | Cosavirus | Enteric | - | + | (Razizadeh et al., 2022) |
| | Enterovirus | Enteric | + | + | (Wells and Coyne, 2019) |
| | Salivirus | Enteric | + | + | (Yu et al., 2015) |
| Pneumoviridae | Orthopneumovirus | Respiratory | + | + | (Rima et al., 2017) |
| | Unclassified Pneumoviridae | Unclassified | + | - | - |
| Polyomaviridae | Betapolyomavirus | Other | + | + | (Prado et al., 2018) |
| | Alphapolyomavirus | Other | + | + | |

| | | | | | |
|---|---|---|---|---|---|
| | Deltapolyomavirus | Other | + | + | |
| Poxviridae | Orthopoxvirus | Respiratory, Other | - | + | (Pauli et al., 2010) |
| | Molluscipoxvirus | Other | + | + | (Tyring, 2003) |
| Reoviridae | Rotavirus | Enteric | - | + | (Crawford et al., 2017) |
| | Orthoreovirus | Respiratory, Enteric | - | + | (Chua et al., 2011) |
| Retroviridae | Lentivirus | Bloodborne | + | + | (Deeks et al., 2015) |
| Smacoviridae | Huchismacovirus | Unknown | + | + | (Ng et al., 2015) |
| Tobaniviridae | Torovirus | Enteric | - | + | (Ujike and Taguchi, 2021) |

**References**


Bosch, A., Pintó, R.M., Guix, S., 2014. Human astroviruses. Clin Microbiol Rev 27, 1048–1074. https://doi.org/10.1128/CMR.00013-14

Chua, K.B., Voon, K., Yu, M., Keniscope, C., Abdul Rasid, K., Wang, L.F., 2011. Investigation of a potential zoonotic transmission of orthoreovirus associated with acute influenza-like illness in an adult patient. PLoS One 6. https://doi.org/10.1371/journal.pone.0025434

Crawford, S.E., Ramani, S., Tate, J.E., Parashar, U.D., Svensson, L., Hagbom, M., Franco, M.A., Greenberg, H.B., O'Ryan, M., Kang, G., Desselberger, U., Estes, M.K., 2017. Rotavirus infection. Nat Rev Dis Primers 3. https://doi.org/10.1038/nrdp.2017.83

de Bolle, L., Naesens, L., de Clercq, E., 2005. Update on human herpesvirus 6 biology, clinical features, and therapy. Clin Microbiol Rev 18, 217–245. https://doi.org/10.1128/CMR.18.1.217-245.2005

de Crom, S.C.M., Rossen, J.W.A., van Furth, A.M., Obihara, C.C., 2016. Enterovirus and parechovirus infection in children: a brief overview. Eur J Pediatr 175, 1023–1029. https://doi.org/10.1007/s00431-016-2725-7

Deeks, S.G., Overbaugh, J., Phillips, A., Buchbinder, S., 2015. HIV infection. Nat Rev Dis Primers 1. https://doi.org/10.1038/nrdp.2015.35

Dioverti, M.V., Razonable, R.R., 2016. Cytomegalovirus. Microbiol Spectr 4. https://doi.org/10.1128/microbiolspec.DMIH2-0022-2015

Doorbar, J., Egawa, N., Griffin, H., Kranjec, C., Murakami, I., 2015. Human papillomavirus molecular biology and disease association. Rev Med Virol. https://doi.org/10.1002/rmv.1822

Ganesh, B., Bányai, K., Martella, V., Jakab, F., Masachessi, G., Kobayashi, N., 2012. Picobirnavirus infections: Viral persistence and zoonotic potential. Rev Med Virol 22, 245–256. https://doi.org/10.1002/rmv.1707

Ghebremedhin, B., 2014. Human adenovirus: Viral pathogen with increasing importance. Eur J Microbiol Immunol (Bp) 4, 26–33. https://doi.org/10.1556/eujmi.4.2014.1.2

Glass, R.I., Parashar, U.D., Estes, M.K., 2009. Norovirus gastroenteritis. New England Journal of Medicine. https://doi.org/10.1056/NEJMra0804575

Jager, M.C., Tomlinson, J.E., Lopez-Astacio, R.A., Parrish, C.R., van de Walle, G.R., 2021. Small but mighty: old and new parvoviruses of veterinary significance. Virol J 18. https://doi.org/10.1186/s12985-021-01677-y

Kaczorowska, J., van der Hoek, L., 2020. Human anelloviruses: Diverse, omnipresent and commensal members of the virome. FEMS Microbiol Rev 44, 305–313. https://doi.org/10.1093/femsre/fuaa007

Khamrin, P., Maneekarn, N., Okitsu, S., Ushijima, H., 2014. Epidemiology of human and animal kobuviruses. Virusdisease 25, 195–200. https://doi.org/10.1007/s13337-014-0200-5

Kutter, J.S., Spronken, M.I., Fraaij, P.L., Fouchier, R.A., Herfst, S., 2018. Transmission routes of respiratory viruses among humans. Curr Opin Virol. https://doi.org/10.1016/j.coviro.2018.01.001

Lemon, S.M., Ott, J.J., van Damme, P., Shouval, D., 2018. Type A viral hepatitis: A summary and update on the molecular virology, epidemiology, pathogenesis and prevention. J Hepatol 68, 167–184. https://doi.org/10.1016/j.jhep.2017.08.034

Li, L., Kapoor, A., Slikas, B., Bamidele, O.S., Wang, C., Shaukat, S., Masroor, M.A., Wilson, M.L., Ndjango, J.-B.N., Peeters, M., Gross-Camp, N.D., Muller, M.N., Hahn, B.H., Wolfe, N.D., Triki, H., Bartkus, J., Zaidi, S.Z., Delwart, E., 2010. Multiple Diverse Circoviruses Infect Farm Animals and Are Commonly Found in Human and Chimpanzee Feces. J Virol 84, 1674–1682. https://doi.org/10.1128/jvi.02109-09

McBride, A.A., 2022. Human papillomaviruses: diversity, infection and host interactions. Nat Rev Microbiol 20, 95–108. https://doi.org/10.1038/s41579-021-00617-5

Mesri, E.A., Cesarman, E., Boshoff, C., 2010. Kaposi's sarcoma herpesvirus/ Human herpesvirus-8 (KSHV/HHV8), and the oncogenesis of Kaposi's sarcoma. Nat Rev Cancer 10, 707–719. https://doi.org/10.1038/nrc2888

Ng, T.F.F., Zhang, W., Sachsenrö Der, J., Kondov, N.O., da Costa, A.C., Vega, E., Holtz, L.R., Wu, G., Wang, D., Stine, C.O., Antonio, M., Mulvaney, U.S., Muench, M.O., Deng, X., Ambert-Balay, K., Pothier, P., Vinjé, J., Delwart, E., 2015. A diverse group of small circular ssDNA viral genomes in human and non-human primate stools. Virus Evol 1. https://doi.org/10.1093/ve/vev017

Oka, T., Wang, Q., Katayama, K., Saifb, L.J., 2015. Comprehensive review of human sapoviruses. Clin Microbiol Rev 28, 32–53. https://doi.org/10.1128/CMR.00011-14

Pauli, G., Blümel, J., Burger, R., Drosten, C., Gröner, A., Gürtler, L., Heiden, M., Hildebrandt, M., Jansen, B., Montag-Lessing, T., Offergeld, R., 2010. Orthopox viruses: Infections in humans. Transfusion Medicine and Hemotherapy 37, 351–364. https://doi.org/10.1159/000322101

Prado, J.C.M., Monezi, T.A., Amorim, A.T., Lino, V., Paladino, A., Boccardo, E., 2018. Human polyomaviruses and cancer: An overview. Clinics 73. https://doi.org/10.6061/clinics/2018/e558s

Qiu, J., Söderlund-Venermo, M., Young, N.S., 2017. Human parvoviruses. Clin Microbiol Rev 30, 43–113. https://doi.org/10.1128/CMR.00040-16

Razizadeh, M.H., Khatami, A., Zarei, M., 2022. Global Status of Bufavirus, Cosavirus, and Saffold Virus in Gastroenteritis: A Systematic Review and Meta-Analysis. Front Med (Lausanne) 8. https://doi.org/10.3389/fmed.2021.775698

Rima, B., Balkema-Buschmann, A., Dundon, W.G., Duprex, P., Easton, A., Fouchier, R., Kurath, G., Lamb, R., Lee, B., Rota, P., Wang, L., Ictv Report Consortium, 2019. ICTV Virus Taxonomy Profile: Paramyxoviridae. J Gen Virol 100, 1593–1594. https://doi.org/10.1099/jgv.0.001328

Rima, B., Collins, P., Easton, A., Fouchier, R., Kurath, G., Lamb, R.A., Lee, B., Maisner, A., Rota, P., Wang, L., 2017. ICTV virus taxonomy profile: Pneumoviridae. Journal of General Virology 98, 2912–2913. https://doi.org/10.1099/jgv.0.000959

Smith, J.M., Uvin, A.Z., Macmadu, A., Rich, J.D., 2017. Epidemiology and Treatment of Hepatitis B in Prisoners. Curr Hepatol Rep 16, 178–183. https://doi.org/10.1007/s11901-017-0364-8

Tsao, S.W., Tsang, C.M., To, K.F., Lo, K.W., 2015. The role of Epstein-Barr virus in epithelial malignancies. Journal of Pathology 235, 323–333. https://doi.org/10.1002/path.4448

Tyring, S.K., 2003. Molluscum contagiosum: The importance of early diagnosis and treatment. Am J Obstet Gynecol 189. https://doi.org/10.1067/S0002-9378(03)00793-2

Ujike, M., Taguchi, F., 2021. Recent progress in torovirus molecular biology. Viruses 13. https://doi.org/10.3390/v13030435

Wang, D., Urisman, A., Liu, Y.T., Springer, M., Ksiazek, T.G., Erdman, D.D., Mardis, E.R., Hickenbotham, M., Magrini, V., Eldred, J., Latreille, J.P., Wilson, R.K., Ganem, D., DeRisi, J.L., 2003. Viral discovery and sequence recovery using DNA microarrays. PLoS Biol 1. https://doi.org/10.1371/journal.pbio.0000002

Wells, A.I., Coyne, C.B., 2019. Enteroviruses: A gut-wrenching game of entry, detection, and evasion. Viruses 11, 1–20. https://doi.org/10.3390/v11050460

Widener, R.W., Whitley, R.J., 2014. Herpes simplex virus, 1st ed, Handbook of Clinical Neurology. Elsevier B.V. https://doi.org/10.1016/B978-0-444-53488-0.00011-0

Yu, J.M., Ao, Y.Y., Liu, N., Li, L.L., Duan, Z.J., 2015. Salivirus in children and its association with childhood acute gastroenteritis: A paired case-control study. PLoS One 10. https://doi.org/10.1371/journal.pone.0130977
